# Neighbourhood-level burden of social risk factors on respiratory syncytial virus hospitalization in Ontario, Canada, 2016-2019

**DOI:** 10.1101/2024.02.27.24303436

**Authors:** Kitty Y.A. Chen, Trevor van Ingen, Brendan T. Smith, Tiffany Fitzpatrick, Michael Whelan, Alyssa S. Parpia, Jenna Alessandrini, Sarah A. Buchan

## Abstract

**Introduction:** Beyond clinical risk factors, little is known about the impact of social determinants on respiratory syncytial virus (RSV) burden. Our study aimed to estimate RSV-related hospitalization rates across sociodemographic and housing characteristics.

**Methods:** We conducted a population-based study of all RSV-related hospitalizations in Ontario, Canada between September 1, 2016 and August 31, 2019 using validated hospital discharge codes and census data. Crude and age-standardized annualized RSV incidence rates and rate ratios (RR) were estimated for a range of individual-level demographics and neighbourhood-level measures of marginalization and housing characteristics.

**Results:** Overall, the annual RSV-related hospitalization rate was 27 per 100,000, with the highest rates observed in children <12 months (1049 per 100,000) and 12-23 months (294 per 100,000), and adults ≥85 years (155 per 100,000). Higher RSV-related hospitalization rates were associated with increasing marginalization quintile (Q) of material resources (RR: 1.4; Q5: 33 per 100,000 versus Q1: 24 per 100,000) and household instability (RR: 1.5; Q5: 31 per 100,000 versus Q1: 22 per 100,000).

**Conclusion:** The burden of RSV-related hospitalization was greatest in young children and older adults, with variation by sociodemographic and housing factors. Understanding the role of these social factors is crucial for informing equitable preventive program delivery.

## Introduction

Respiratory syncytial virus (RSV) is a leading cause of lower respiratory tract infection among young children, older adults, and immunocompromised individuals [1]. RSV infection represents a large burden on the healthcare system on a global scale, including within Canada, contributing to approximately 24.8 million acute respiratory infection episodes and 76,600 deaths globally each year [1–3]. Substantial progress has been made in the field of RSV vaccines and immunoprophylaxis, which will help mitigate RSV-related hospitalization and the burden on the healthcare system; however, ensuring equitable access and widespread uptake of these products across population groups will be crucial for their success. This dynamic landscape, coupled with the emergence of RSV vaccines and new monoclonal antibodies for different age groups, underscores the importance of reliable RSV-related hospitalization estimates to support health system preparedness and cost-effective delivery of targeted interventions [4]. Little is known about the baseline risk in groups with high RSV burden, especially in the Canadian context.

While clinical risk factors for severe RSV disease are well-recognized, it is crucial to understand the role that social and environmental variables play in severe RSV disease burden. Social factors are major determinants of morbidity and mortality, yielding distinct differences in inpatient medical services. In the context of infectious diseases, the COVID-19 pandemic further emphasized the importance of understanding the interplay between infectious diseases and social determinants of health, at both individual and neighbourhood levels [5–8]. Social and structural inequities can contribute to unjust differences in how individuals are affected by RSV, such as susceptibility, contact patterns, adoption of preventive measures, and access to quality treatment [6].

Despite the variable dynamics in the timing and magnitude of RSV epidemics across communities, little is known about the differential burdens of RSV-related hospitalizations by socioeconomic status [9]. Experiences of RSV may vary by region due to shared socioeconomic characteristics among community members [10]. Additionally, cohort studies of RSV infection have revealed that the social structuring and size of households play an important role in RSV transmission dynamics, in terms of characterizing contact patterns within households [11–13]. Moreover, there is a lack of comprehensive investigation into the social risk factors associated with RSV-related hospitalizations across the age spectrum beyond infancy [14,15]. As such, the purpose of our study was to provide age-specific estimates of RSV-related hospitalization in Ontario, Canada, with a focus on understanding trends by individual-level demographics, and neighbourhood-level sociodemographic and housing characteristics.

## Methods

### Study population

We examined individuals in Ontario, Canada with an eligible RSV-related hospitalization from September 1, 2016, to August 31, 2019. An RSV season was defined as September 1^st^ to August 31^st^ of the following year. Of 277,852 hospital admittances for acute respiratory infections from September 1, 2016, to August 31, 2019, 11,746 hospitalizations had an RSV-coded diagnosis. We excluded patients without a recorded provincial health insurance number, benefiting from Canada’s universal healthcare coverage of basic medical and emergency services resulting in 11,655 hospitalizations. After accounting for acute hospital transfers, there were a total of 11,311 hospitalizations. If repeated RSV-related hospitalizations occurred within 30 days of one another and between seasons, subsequent admissions were excluded from the study to prevent the same infection from being captured more than once, resulting in 11,305 hospitalizations. For individuals with more than one RSV-related hospitalization in a given season, we included the earliest hospitalization for each season after accounting for any readmissions as previously described [16,17]. A total of 11,039 RSV-related hospitalizations were included in the study. Further inclusion and exclusion criteria are described in Figure S1.

### Data source

Information on all inpatient RSV-related hospitalizations and their demographics were obtained from the Canadian Institute of Health Information (CIHI) Discharge Abstract Database (DAD). Neighbourhood-level sociodemographic and housing characteristics were obtained from the 2016 Canadian census provided by Statistics Canada and the 2016 census-based Ontario Marginalization (ON-Marg) index. The 2011 Canadian census age structure was used to conduct age standardization.

### Outcomes

RSV-related hospitalizations were defined as a hospitalization with at least one RSV-related International Classification of Diseases, Tenth Revision, Canada (ICD-10-CA) code: RSV pneumonia (J12.1), acute bronchitis due to RSV (J20.5), acute bronchiolitis due to RSV (J21.0), and RSV as the cause of disease classified elsewhere (B97.4) in any of the all diagnosis fields. Prior validation studies have shown these codes to have extremely high specificity (>99%) for identifying RSV-related admissions in both pediatric and adult populations in Ontario, Canada compared to laboratory-confirmed admissions, with sensitivity estimates ranging from moderate (69%) to high (>99%), in the general and pediatric populations, respectively [18,19]. We also examined intensive care unit (ICU) admissions, in-hospital deaths and their associated median length of stay were also examined. Length of stay was calculated using the earliest admission date and latest discharge date, including inpatient transfers. Additionally, we examined RSV as the most responsible diagnosis, which is the diagnosis that responsible for the patient’s greatest length of stay or resource utilization [20]. If inpatient transfers occurred, the earliest most responsible diagnosis in the first transfer was considered.

### Individual and neighbourhood-level exposures

Age at the time of admission, sex, and month of hospital admission were examined at the individual level. Census-based neighbourhood-level measures were used to ascertain housing characteristics; these were measured at the dissemination area (DA) level, Canada’s smallest census area unit of 400 to 700 persons [21]. Census-based housing characteristics included multi-generational families, household crowding, average dwelling size and households with persons under 5 years old. Multi-generational households represented households with at least one person who is both the child of a person in the household and a grandchild to another person in the same household [22]. Household crowding was defined as having more than one person per room in a private dwelling [23]. Unsuitable housing referred to having fewer bedrooms for the size and composition of the household, according to the National Occupancy Standard [24]. Ontario’s 34 public health units were categorized accordingly into seven routinely used aggregated public health regions [25]. Rurality was defined using the postal code Forward Sortation Area [26].

We also assessed census-based constructs included in the 2016 ON-Marg index, which measures area-level marginalization across four dimensions: racialized and newcomer populations, material resources, households and dwellings, as well as age and labour force [21,27]. The racialized and newcomer populations dimension measured the proportion of recent immigrants and the proportion of the population who self-identify as a visible minority living within an area. Lack of material resources represented poverty and the inability to purchase basic material needs. The households and dwellings dimension was related to residential characteristics and certain familial structure characteristics that have not been previously mentioned such as residential mobility, ownership and occupancy. Age and labour force captured age structure characteristics and the proportion of individuals not participating in the labour force [21,27].

We used the ON-Marg index to assign each individual to a level of marginalization based on their lived neighbourhood [21,27]. Cases were assigned to DAs based on postal code of residence using the Postal Code Conversion File (PCCF) Plus version 7E [28]. ON-Marg index values and census indicators were categorized into quintiles, derived by ranking all Ontario DAs on their ON-Marg or census indicator values and categorizing them into five equal groups, ordered from Quintile (Q) 1 (e.g., least marginalized/least multi-generational, etc.) to Q5 (most marginalized/most multi-generational, etc.).

### Statistical analysis

Hospitalization rates and rate ratios (RRs), along with associated 95% confidence intervals (CIs), were calculated overall, and by demographic, sociodemographic, and housing characteristics.

Age-specific rates were calculated for RSV-related hospitalizations among those aged <12 months, 12-23 months, 2-4 years, 5-17 years, 18-49 years, 50-64 years, and ≥65 years. RSV-related hospitalization rates for sociodemographic and housing characteristics were stratified by broader age groups at admission (<5, 5-64, ≥65 years) to account for the larger number of cases in young children and older adults. Annualized hospitalization incidence was calculated using the cumulative number of RSV-related hospitalizations as the numerator and the 2016 Ontario portion of the Canadian census, multiplied by three to account for the three RSV seasons, as the denominator. Rates for the sociodemographic and housing characteristics were age-standardized using the 2011 Ontario portion of the Canadian census by <5, 5-64, and ≥65 years of age to account for age differences between quintiles. Demographic, sociodemographic and housing characteristics were further stratified by ICU admission and death in hospital, following RSV-related hospitalization. Additionally, the cumulative proportion of RSV-related hospitalization by month was stratified by age to observe growth in cases.

We conducted sensitivity analyses with RSV as the most responsible diagnosis. Further, we examined the top five most responsible diagnoses of patients without RSV as their most responsible diagnosis by age.

We obtained ethical approval from Public Health Ontario’s Research Ethics Board. Statistical analyses were performed in SAS version 9.4.

## Results

### Study population

A total of 11,039 RSV-related hospitalizations were included in the three-year study period, with most of these occurring among patients aged <12 months (38.5%), those aged ≥65 years (31.5%) and those aged 12-23 months (10.9%), as illustrated in **Table 1**. RSV-related hospitalizations most frequently occurred during winter months (December [25.5%], January [30.6%], and February [17.8%]). The cumulative proportion of RSV-related hospitalizations rose fastest in those aged 2-4 years, aged 12-23 months, followed by <12 months (**Figure S2**).

**Table 1:**
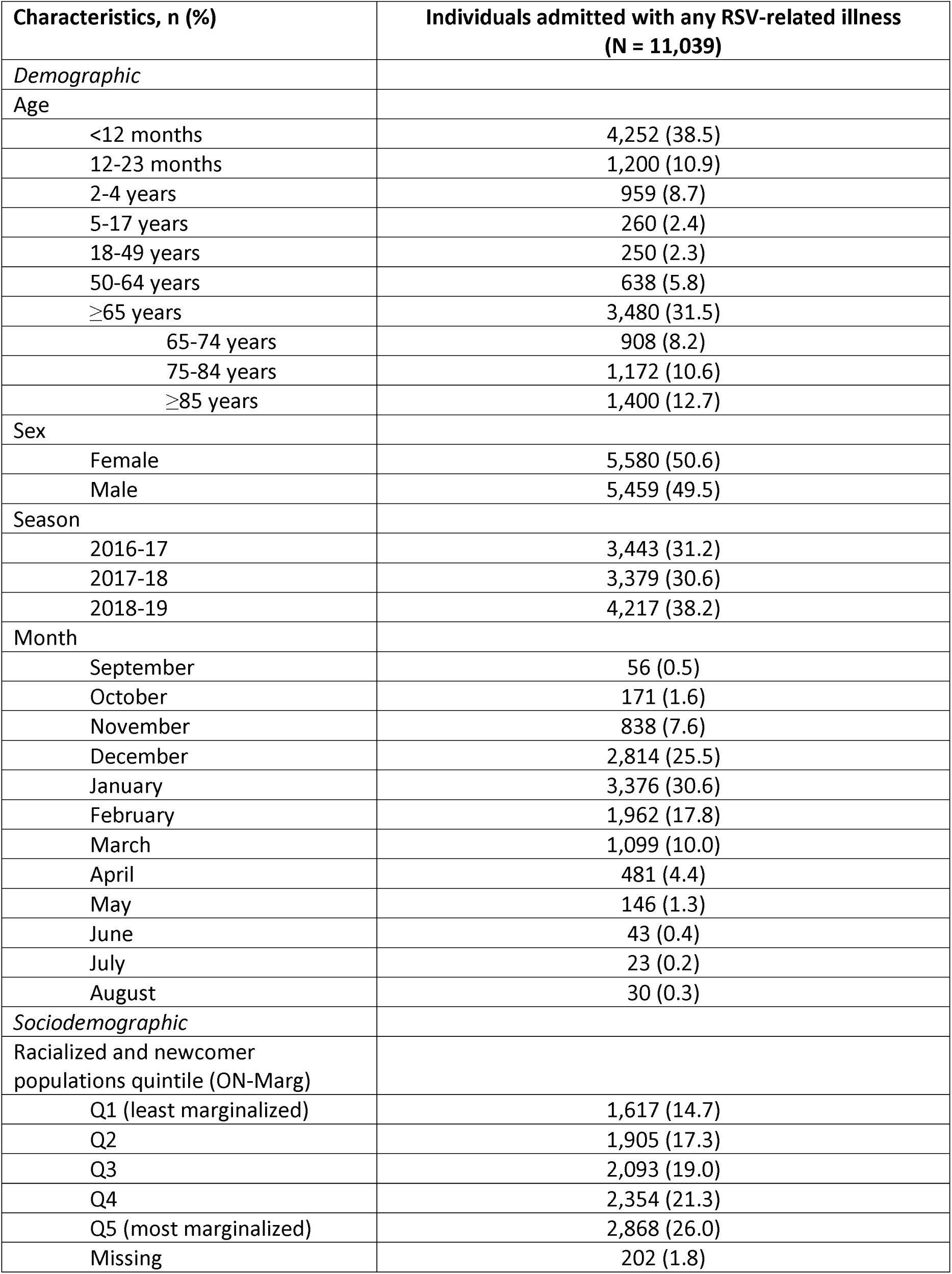
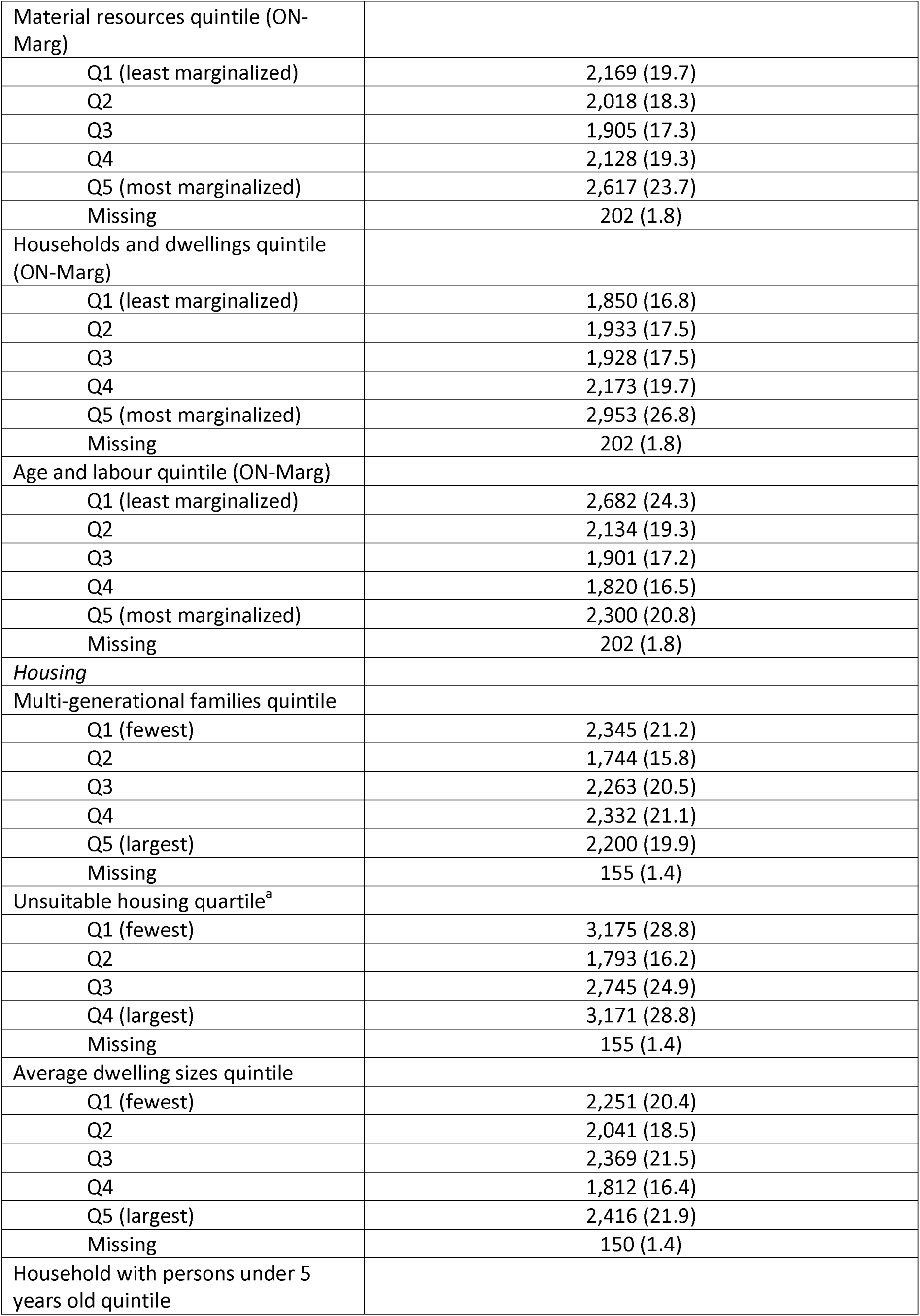
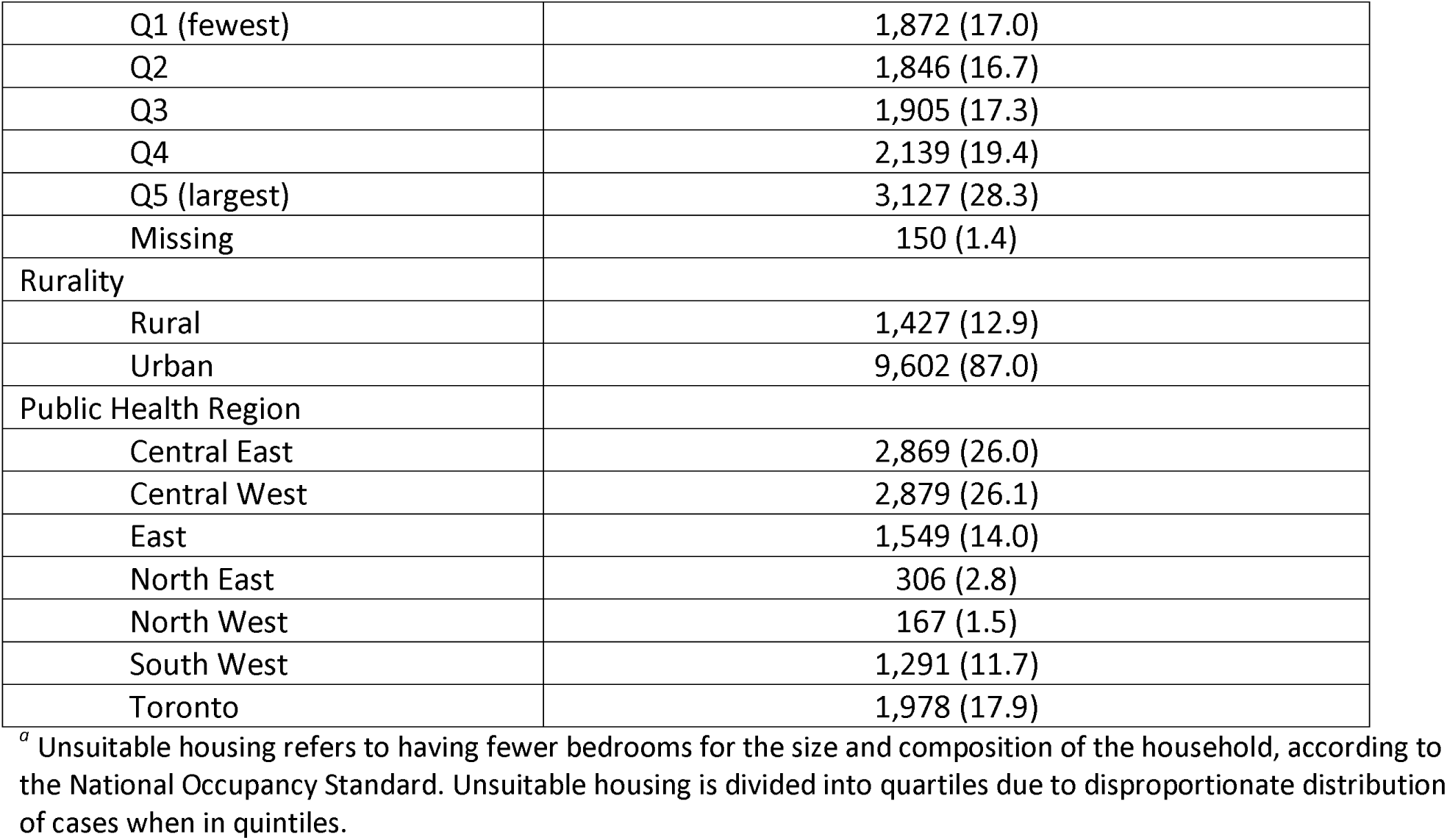
Characteristics of patients hospitalized for an RSV-related illness in Ontario from September 1, 2016 to August 31, 2019.

| Characteristics, n (%) | Individuals admitted with any RSV-related illness<br>(N = 11,039) |
| --- | --- |
| *Demographic* |  |
| Age |  |
| <12 months | 4,252 (38.5) |
| 12-23 months | 1,200 (10.9) |
| 2-4 years | 959 (8.7) |
| 5-17 years | 260 (2.4) |
| 18-49 years | 250 (2.3) |
| 50-64 years | 638 (5.8) |
| ≥65 years | 3,480 (31.5) |
| 65-74 years | 908 (8.2) |
| 75-84 years | 1,172 (10.6) |
| ≥85 years | 1,400 (12.7) |
| Sex |  |
| Female | 5,580 (50.6) |
| Male | 5,459 (49.5) |
| Season |  |
| 2016-17 | 3,443 (31.2) |
| 2017-18 | 3,379 (30.6) |
| 2018-19 | 4,217 (38.2) |
| Month |  |
| September | 56 (0.5) |
| October | 171 (1.6) |
| November | 838 (7.6) |
| December | 2,814 (25.5) |
| January | 3,376 (30.6) |
| February | 1,962 (17.8) |
| March | 1,099 (10.0) |
| April | 481 (4.4) |
| May | 146 (1.3) |
| June | 43 (0.4) |
| July | 23 (0.2) |
| August | 30 (0.3) |
| *Sociodemographic* |  |
| Racialized and newcomer<br>populations quintile (ON-Marg) |  |
| Q1 (least marginalized) | 1,617 (14.7) |
| Q2 | 1,905 (17.3) |
| Q3 | 2,093 (19.0) |
| Q4 | 2,354 (21.3) |
| Q5 (most marginalized) | 2,868 (26.0) |
| Missing | 202 (1.8) |
| Material resources quintile (ON-Marg) |  |
| Q1 (least marginalized) | 2,169 (19.7) |
| Q2 | 2,018 (18.3) |
| Q3 | 1,905 (17.3) |
| Q4 | 2,128 (19.3) |
| Q5 (most marginalized) | 2,617 (23.7) |
| Missing | 202 (1.8) |
| Households and dwellings quintile (ON-Marg) |  |
| Q1 (least marginalized) | 1,850 (16.8) |
| Q2 | 1,933 (17.5) |
| Q3 | 1,928 (17.5) |
| Q4 | 2,173 (19.7) |
| Q5 (most marginalized) | 2,953 (26.8) |
| Missing | 202 (1.8) |
| Age and labour quintile (ON-Marg) |  |
| Q1 (least marginalized) | 2,682 (24.3) |
| Q2 | 2,134 (19.3) |
| Q3 | 1,901 (17.2) |
| Q4 | 1,820 (16.5) |
| Q5 (most marginalized) | 2,300 (20.8) |
| Missing | 202 (1.8) |
| *Housing* |  |
| Multi-generational families quintile |  |
| Q1 (fewest) | 2,345 (21.2) |
| Q2 | 1,744 (15.8) |
| Q3 | 2,263 (20.5) |
| Q4 | 2,332 (21.1) |
| Q5 (largest) | 2,200 (19.9) |
| Missing | 155 (1.4) |
| Unsuitable housing quartile<sup>a</sup> |  |
| Q1 (fewest) | 3,175 (28.8) |
| Q2 | 1,793 (16.2) |
| Q3 | 2,745 (24.9) |
| Q4 (largest) | 3,171 (28.8) |
| Missing | 155 (1.4) |
| Average dwelling sizes quintile |  |
| Q1 (fewest) | 2,251 (20.4) |
| Q2 | 2,041 (18.5) |
| Q3 | 2,369 (21.5) |
| Q4 | 1,812 (16.4) |
| Q5 (largest) | 2,416 (21.9) |
| Missing | 150 (1.4) |
| Household with persons under 5 years old quintile |  |
| Q1 (fewest) | 1,872 (17.0) |
| Q2 | 1,846 (16.7) |
| Q3 | 1,905 (17.3) |
| Q4 | 2,139 (19.4) |
| Q5 (largest) | 3,127 (28.3) |
| Missing | 150 (1.4) |
| Rurality |  |
| Rural | 1,427 (12.9) |
| Urban | 9,602 (87.0) |
| Public Health Region |  |
| Central East | 2,869 (26.0) |
| Central West | 2,879 (26.1) |
| East | 1,549 (14.0) |
| North East | 306 (2.8) |
| North West | 167 (1.5) |
| South West | 1,291 (11.7) |
| Toronto | 1,978 (17.9) |

<sup>a</sup> Unsuitable housing refers to having fewer bedrooms for the size and composition of the household, according to the National Occupancy Standard. Unsuitable housing is divided into quartiles due to disproportionate distribution of cases when in quintiles.

### Age-specific patterns of annual and monthly RSV-related hospitalization rate

Throughout the study period, the rate of RSV-related hospitalizations exhibited clear seasonality increasing from September to a peak in January across the three seasons before declining again to lows in July and August (**Figure 1**). The highest rates were consistently seen among patients aged <12 months, followed by those aged 12-23 months, 2-4 years, and ≥65 years.

**Figure 1:**
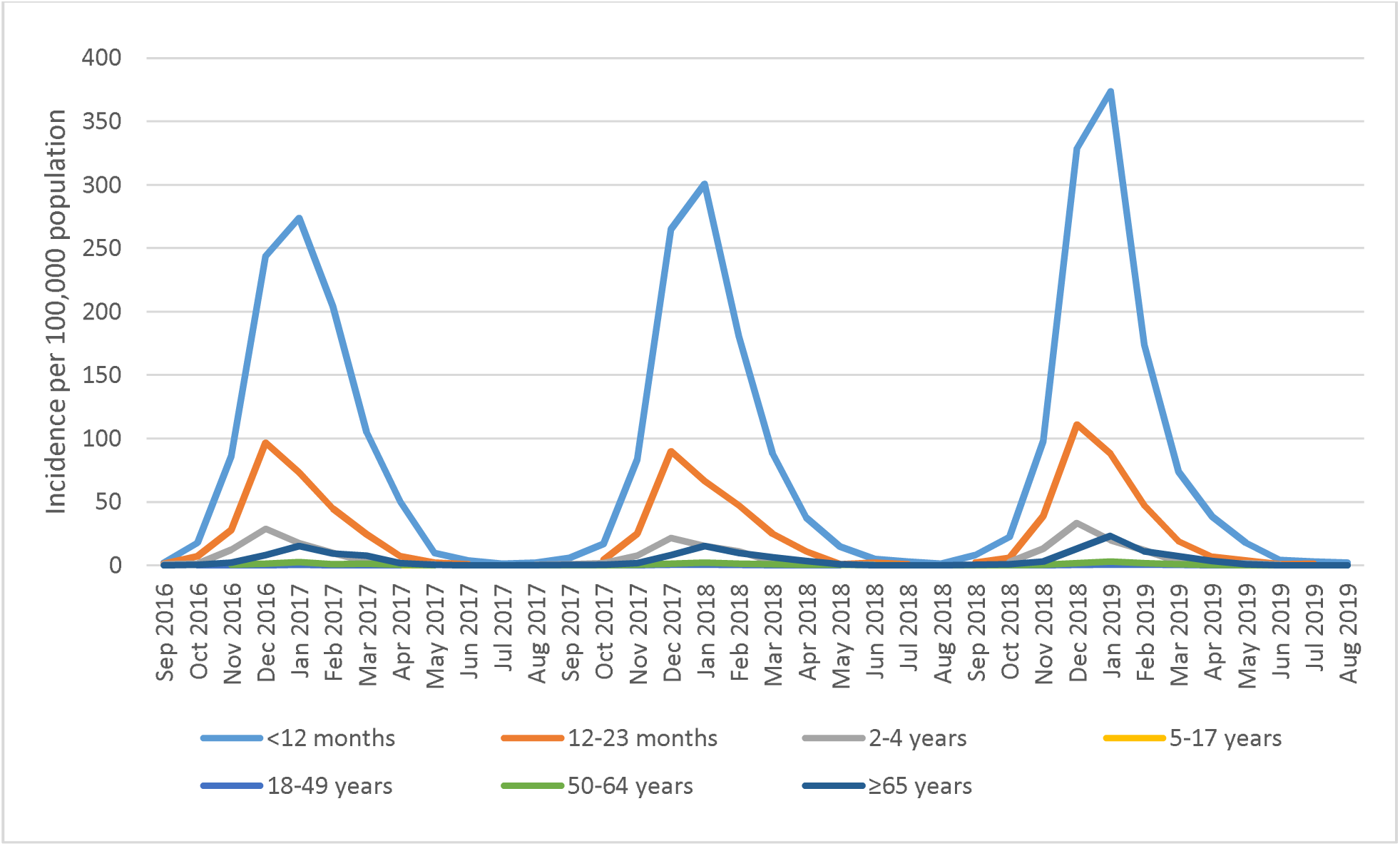
Age-specific monthly RSV-related hospitalization rates, September 1, 2016 to August 31, 2019.

### Age-standardized patterns of RSV-related hospitalization rate

The highest annual incidence of hospitalizations attributable to RSV infections was found in patients <12 months [927.8 per 100,000; 95% CI (898.2, 957.5)], followed by those 12-23 months [294.4 per 100,000; 95% (277.7, 311.0)], and ≥85 years [155.1 per 100,000; 95% CI (147.0, 163.2)] (**Table 2A**). No differences in RSV-related hospitalization rates were observed by sex. Overall, the age-standardized RSV-related hospitalization rate was 27.3 cases per 100,000, which varied across sociodemographic characteristics (**Table 2B**). There was no clear gradient in RSV-related hospitalizations across quintiles of the racialized and newcomer populations index. We observed an increase in RSV-related hospitalization associated with increasing material resource-related marginalization (32.6 per 100,000 versus 24.1 per 100,000) (**Figure 2**). In terms of households and dwellings dimension, neighbourhoods with the most households and dwellings-related marginalization exhibited higher rates of hospitalization compared to the least marginalized neighbourhoods (31.4 per 100,000 versus 21.5 per 100,000). Increasing age and labour force-related marginalization was associated with an increase in RSV-related hospitalization rates (30.9 per 100,000 versus 24.9 per 100,000).

**Table 2A:** Crude annualized RSV-related hospitalization rates per 100,000 in Ontario by demographic characteristics.

| Characteristic | RSV-related hospitalizations, 2016-2019, n (%) | Ontario population, 2016 | Annualized RSV-related hospitalization rate per 100,000<sup>a</sup> (95% CI) |
| --- | --- | --- | --- |
| *Overall* | 11,039 | 13,448,155 |  |
| *Demographic* |  |  |  |
| Age |  |  |  |
| <12 months | 4,252 (38.5) | 135,080 | 1049.3 (1017.7, 1080.8) |
| 12-23 months | 1,200 (10.9) | 135,875 | 294.4 (277.7, 311.0) |
| 2-4 years | 959 (8.7) | 426,510 | 74.9 (70.2, 79.7) |
| 5-17 years | 260 (2.4) | 1,985,175 | 4.4 (3.8, 4.9) |
| 18-49 years | 250 (2.3) | 5,618,050 | 1.5 (1.3, 1.7) |
| 50-64 years | 638 (5.8) | 2,892,625 | 7.4 (6.8, 7.9) |
| ≥65 years | 3,480 (31.5) | 2,249,625 | 51.6 (49.8, 53.3) |
| 65-74 years | 908 (8.2) | 1,265,375 | 23.9 (22.4, 25.5) |
| 75-84 years | 1,172 (10.6) | 683,330 | 57.2 (53.9, 60.4) |
| ≥85 years | 1,400 (12.7) | 300,920 | 155.1 (147.0, 163.2) |
| Sex |  |  |  |
| Female | 5,580 (50.6) | 6,888,235 | 27.0 (26.3, 27.7) |
| Male | 5,459 (49.5) | 6,554,705 | 27.8 (27.0, 28.5) |
| Season<sup>b</sup> |  |  |  |
| 2016-17 | 3,443 (31.2) | 13,448,155 | 25.6 (24.7, 26.5) |
| 2017-18 | 3,379 (30.6) | 13,448,155 | 25.1 (24.3, 26.0) |
| 2018-19 | 4,217 (38.2) | 13,448,155 | 31.4 (30.4, 32.3) |
| Month |  |  |  |
| September | 56 (0.5) | 13,448,155 | 0.1 (0.1, 0.2) |
| October | 171 (1.6) | 13,448,155 | 0.4 (0.4, 0.5) |
| November | 838 (7.6) | 13,448,155 | 2.1 (1.9, 2.2) |
| December | 2,814 (25.5) | 13,448,155 | 7.0 (6.7, 7.2) |
| January | 3,376 (30.6) | 13,448,155 | 8.4 (8.1, 8.7) |
| February | 1,962 (17.8) | 13,448,155 | 4.9 (4.6, 5.1) |
| March | 1,099 (10.0) | 13,448,155 | 2.7 (2.6, 2.9) |
| April | 481 (4.4) | 13,448,155 | 1.2 (1.1, 1.3) |
| May | 146 (1.3) | 13,448,155 | 0.4 (0.3, 0.4) |
| June | 43 (0.4) | 13,448,155 | 0.1 (0.1, 0.1) |
| July | 23 (0.2) | 13,448,155 | 0.1 (0.0, 0.1) |
| August | 30 (0.3) | 13,448,155 | 0.1 (0.0, 0.1) |

*\*Missing rates were not included in table.*
<sup>a</sup> Annualized rate is calculated using the cumulative number of RSV-related hospitalizations from September 1, 2016 to August 31, 2019 as the numerator and the 2016 Ontario Census multiplied by 3 as the denominator.
<sup>b</sup> Rate for season is calculated using the cumulative number of RSV-related hospitalizations from September 1, 2016 to August 31, 2019 as the numerator and the 2016 Ontario Census as the denominator.

**Table 2B:**
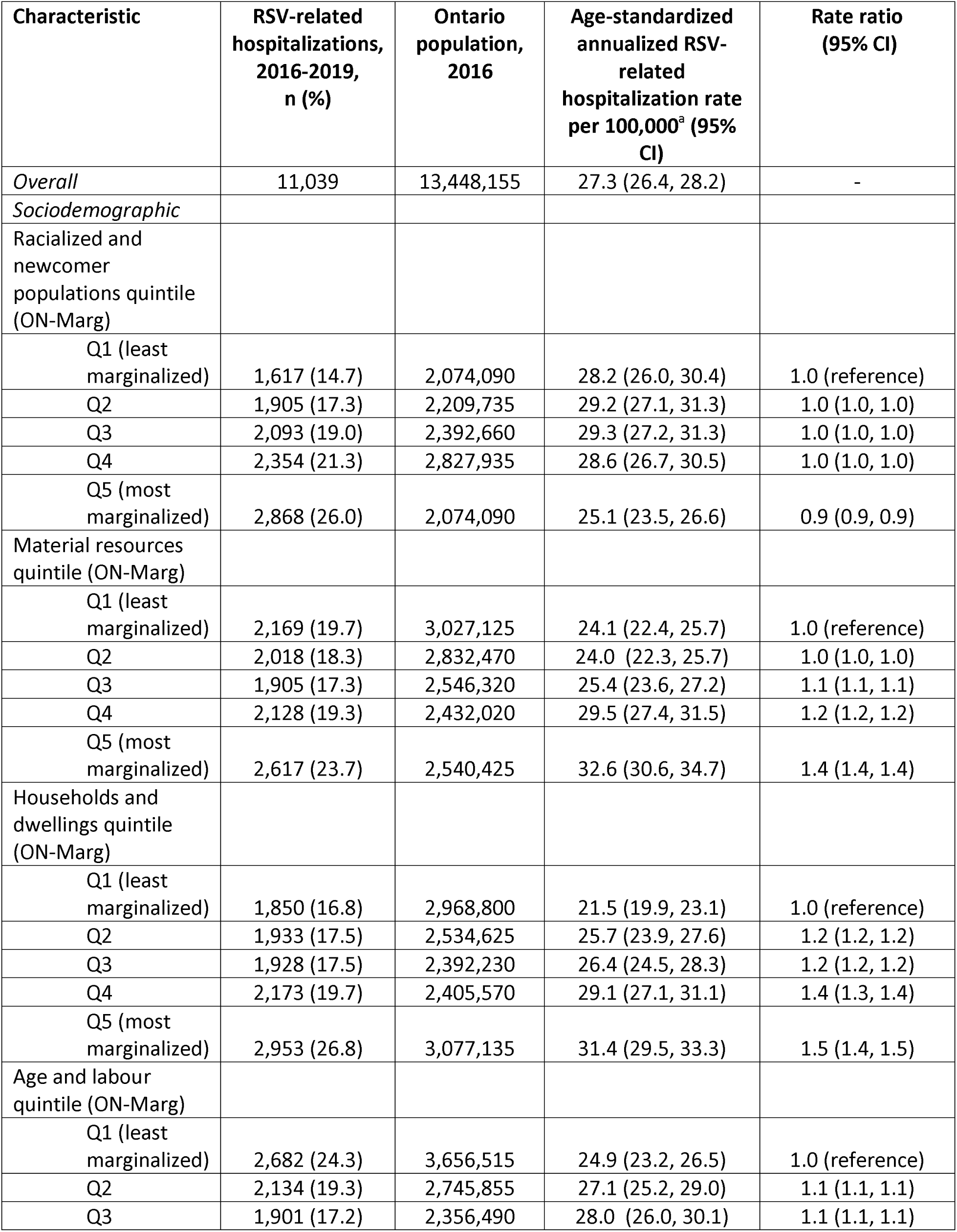
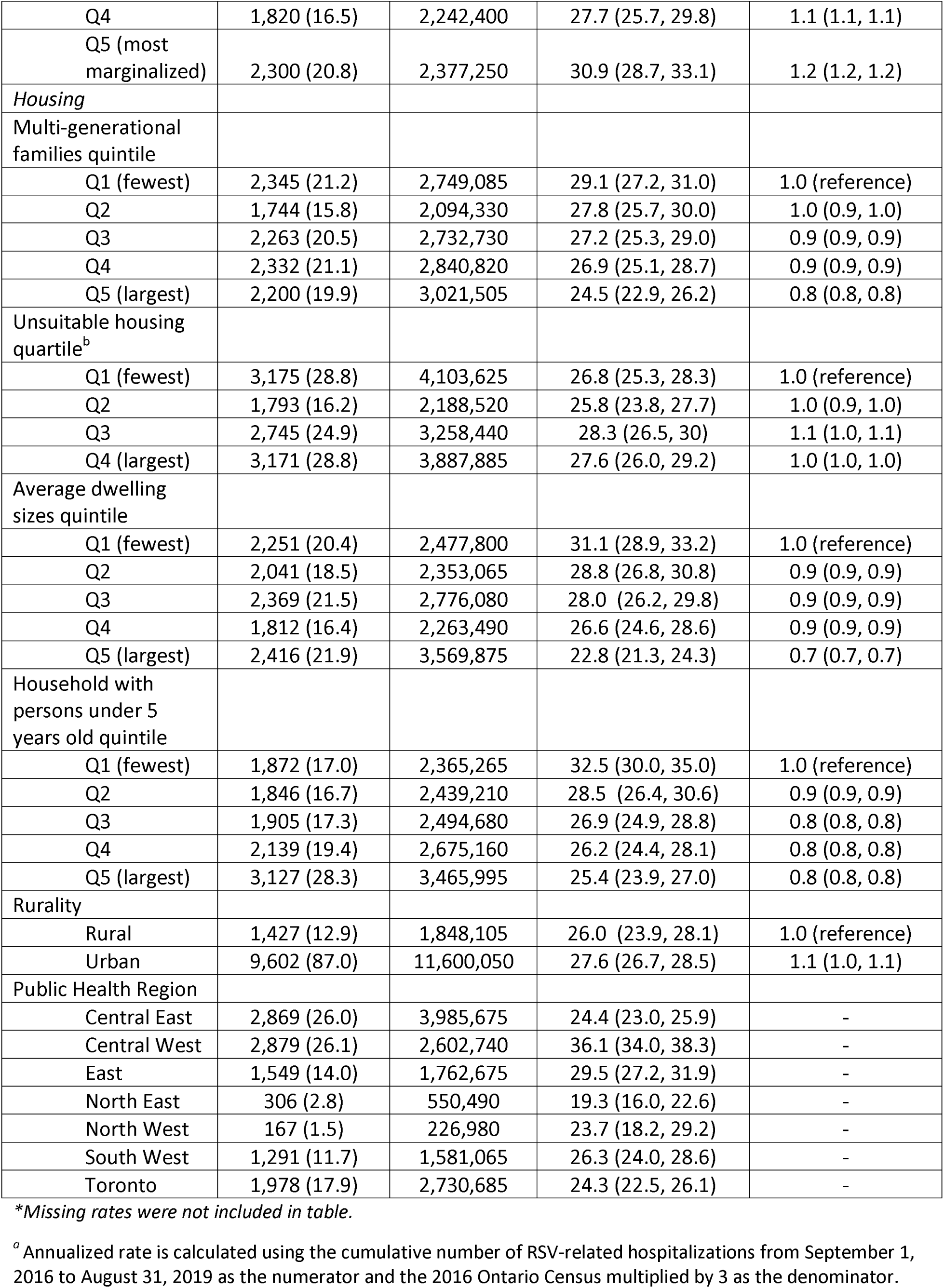
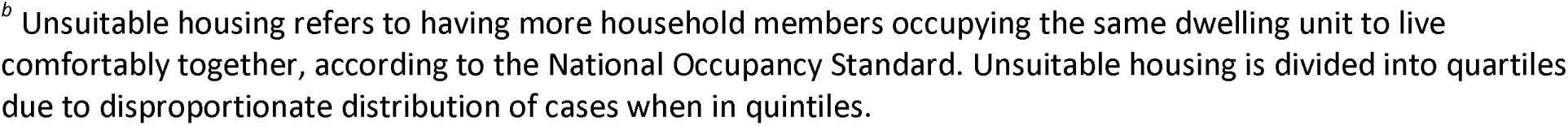
Age-standardized annualized RSV-related hospitalization rates per 100,000 in Ontario by sociodemographic and housing characteristics.

| Characteristic | RSV-related hospitalizations, 2016-2019, n (%) | Ontario population, 2016 | Age-standardized annualized RSV-related hospitalization rate per 100,000<sup>a</sup> (95% CI) | Rate ratio (95% CI) |
| --- | --- | --- | --- | --- |
| *Overall* | 11,039 | 13,448,155 | 27.3 (26.4, 28.2) | - |
| *Sociodemographic* |  |  |  |  |
| Racialized and newcomer populations quintile (ON-Marg) |  |  |  |  |
| Q1 (least marginalized) | 1,617 (14.7) | 2,074,090 | 28.2 (26.0, 30.4) | 1.0 (reference) |
| Q2 | 1,905 (17.3) | 2,209,735 | 29.2 (27.1, 31.3) | 1.0 (1.0, 1.0) |
| Q3 | 2,093 (19.0) | 2,392,660 | 29.3 (27.2, 31.3) | 1.0 (1.0, 1.0) |
| Q4 | 2,354 (21.3) | 2,827,935 | 28.6 (26.7, 30.5) | 1.0 (1.0, 1.0) |
| Q5 (most marginalized) | 2,868 (26.0) | 2,074,090 | 25.1 (23.5, 26.6) | 0.9 (0.9, 0.9) |
| Material resources quintile (ON-Marg) |  |  |  |  |
| Q1 (least marginalized) | 2,169 (19.7) | 3,027,125 | 24.1 (22.4, 25.7) | 1.0 (reference) |
| Q2 | 2,018 (18.3) | 2,832,470 | 24.0 (22.3, 25.7) | 1.0 (1.0, 1.0) |
| Q3 | 1,905 (17.3) | 2,546,320 | 25.4 (23.6, 27.2) | 1.1 (1.1, 1.1) |
| Q4 | 2,128 (19.3) | 2,432,020 | 29.5 (27.4, 31.5) | 1.2 (1.2, 1.2) |
| Q5 (most marginalized) | 2,617 (23.7) | 2,540,425 | 32.6 (30.6, 34.7) | 1.4 (1.4, 1.4) |
| Households and dwellings quintile (ON-Marg) |  |  |  |  |
| Q1 (least marginalized) | 1,850 (16.8) | 2,968,800 | 21.5 (19.9, 23.1) | 1.0 (reference) |
| Q2 | 1,933 (17.5) | 2,534,625 | 25.7 (23.9, 27.6) | 1.2 (1.2, 1.2) |
| Q3 | 1,928 (17.5) | 2,392,230 | 26.4 (24.5, 28.3) | 1.2 (1.2, 1.2) |
| Q4 | 2,173 (19.7) | 2,405,570 | 29.1 (27.1, 31.1) | 1.4 (1.3, 1.4) |
| Q5 (most marginalized) | 2,953 (26.8) | 3,077,135 | 31.4 (29.5, 33.3) | 1.5 (1.4, 1.5) |
| Age and labour quintile (ON-Marg) |  |  |  |  |
| Q1 (least marginalized) | 2,682 (24.3) | 3,656,515 | 24.9 (23.2, 26.5) | 1.0 (reference) |
| Q2 | 2,134 (19.3) | 2,745,855 | 27.1 (25.2, 29.0) | 1.1 (1.1, 1.1) |
| Q3 | 1,901 (17.2) | 2,356,490 | 28.0 (26.0, 30.1) | 1.1 (1.1, 1.1) |
| Q4 | 1,820 (16.5) | 2,242,400 | 27.7 (25.7, 29.8) | 1.1 (1.1, 1.1) |
| Q5 (most marginalized) | 2,300 (20.8) | 2,377,250 | 30.9 (28.7, 33.1) | 1.2 (1.2, 1.2) |
| *Housing* |  |  |  |  |
| Multi-generational families quintile |  |  |  |  |
| Q1 (fewest) | 2,345 (21.2) | 2,749,085 | 29.1 (27.2, 31.0) | 1.0 (reference) |
| Q2 | 1,744 (15.8) | 2,094,330 | 27.8 (25.7, 30.0) | 1.0 (0.9, 1.0) |
| Q3 | 2,263 (20.5) | 2,732,730 | 27.2 (25.3, 29.0) | 0.9 (0.9, 0.9) |
| Q4 | 2,332 (21.1) | 2,840,820 | 26.9 (25.1, 28.7) | 0.9 (0.9, 0.9) |
| Q5 (largest) | 2,200 (19.9) | 3,021,505 | 24.5 (22.9, 26.2) | 0.8 (0.8, 0.8) |
| Unsuitable housing quartile<sup>b</sup> |  |  |  |  |
| Q1 (fewest) | 3,175 (28.8) | 4,103,625 | 26.8 (25.3, 28.3) | 1.0 (reference) |
| Q2 | 1,793 (16.2) | 2,188,520 | 25.8 (23.8, 27.7) | 1.0 (0.9, 1.0) |
| Q3 | 2,745 (24.9) | 3,258,440 | 28.3 (26.5, 30) | 1.1 (1.0, 1.1) |
| Q4 (largest) | 3,171 (28.8) | 3,887,885 | 27.6 (26.0, 29.2) | 1.0 (1.0, 1.0) |
| Average dwelling sizes quintile |  |  |  |  |
| Q1 (fewest) | 2,251 (20.4) | 2,477,800 | 31.1 (28.9, 33.2) | 1.0 (reference) |
| Q2 | 2,041 (18.5) | 2,353,065 | 28.8 (26.8, 30.8) | 0.9 (0.9, 0.9) |
| Q3 | 2,369 (21.5) | 2,776,080 | 28.0 (26.2, 29.8) | 0.9 (0.9, 0.9) |
| Q4 | 1,812 (16.4) | 2,263,490 | 26.6 (24.6, 28.6) | 0.9 (0.9, 0.9) |
| Q5 (largest) | 2,416 (21.9) | 3,569,875 | 22.8 (21.3, 24.3) | 0.7 (0.7, 0.7) |
| Household with persons under 5 years old quintile |  |  |  |  |
| Q1 (fewest) | 1,872 (17.0) | 2,365,265 | 32.5 (30.0, 35.0) | 1.0 (reference) |
| Q2 | 1,846 (16.7) | 2,439,210 | 28.5 (26.4, 30.6) | 0.9 (0.9, 0.9) |
| Q3 | 1,905 (17.3) | 2,494,680 | 26.9 (24.9, 28.8) | 0.8 (0.8, 0.8) |
| Q4 | 2,139 (19.4) | 2,675,160 | 26.2 (24.4, 28.1) | 0.8 (0.8, 0.8) |
| Q5 (largest) | 3,127 (28.3) | 3,465,995 | 25.4 (23.9, 27.0) | 0.8 (0.8, 0.8) |
| *Rurality* |  |  |  |  |
| Rural | 1,427 (12.9) | 1,848,105 | 26.0 (23.9, 28.1) | 1.0 (reference) |
| Urban | 9,602 (87.0) | 11,600,050 | 27.6 (26.7, 28.5) | 1.1 (1.0, 1.1) |
| *Public Health Region* |  |  |  |  |
| Central East | 2,869 (26.0) | 3,985,675 | 24.4 (23.0, 25.9) | - |
| Central West | 2,879 (26.1) | 2,602,740 | 36.1 (34.0, 38.3) | - |
| East | 1,549 (14.0) | 1,762,675 | 29.5 (27.2, 31.9) | - |
| North East | 306 (2.8) | 550,490 | 19.3 (16.0, 22.6) | - |
| North West | 167 (1.5) | 226,980 | 23.7 (18.2, 29.2) | - |
| South West | 1,291 (11.7) | 1,581,065 | 26.3 (24.0, 28.6) | - |
| Toronto | 1,978 (17.9) | 2,730,685 | 24.3 (22.5, 26.1) | - |

*\*Missing rates were not included in table.*
<sup>a</sup> Annualized rate is calculated using the cumulative number of RSV-related hospitalizations from September 1, 2016 to August 31, 2019 as the numerator and the 2016 Ontario Census multiplied by 3 as the denominator.
<sup>b</sup> Unsuitable housing refers to having more household members occupying the same dwelling unit to live comfortably together, according to the National Occupancy Standard. Unsuitable housing is divided into quartiles due to disproportionate distribution of cases when in quintiles.

**Figure 2:**
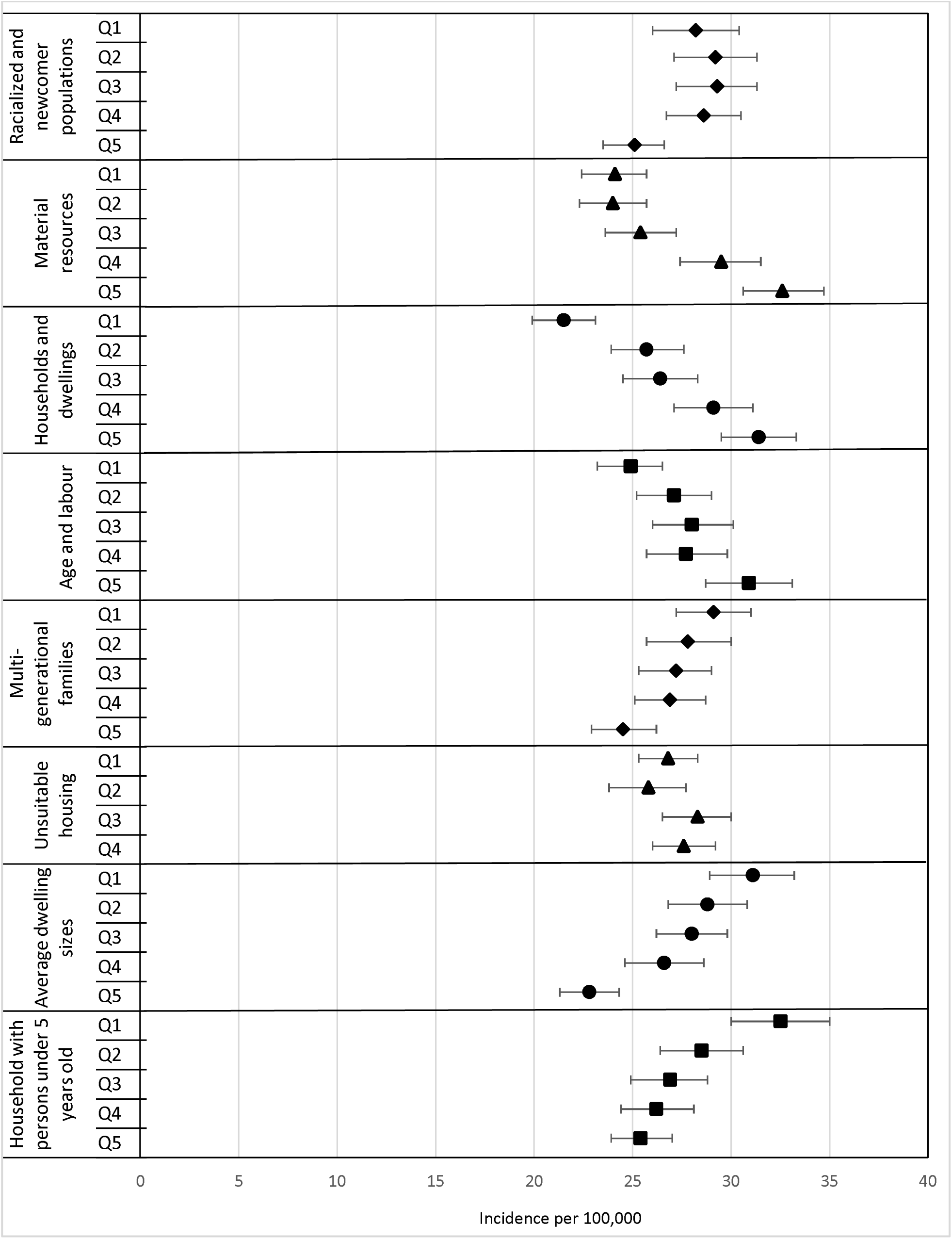
Age-standardized annualized RSV-related hospitalization rates per 100,000 in Ontario, by neighbourhood-level sociodemographic and housing characteristics.

We observed an inverse gradient between many housing characteristics and age-standardized RSV-related hospitalization rates. For example, this was evident in comparing the lowest to highest quintiles of multi-generational families (29.1 per 100,000 versus 24.5 per 100,000), unsuitable housing (31.4 per 100,000 versus 21.5 per 100,000), average dwelling size (31.1 per 100,000 versus 22.8 per 100,000), and households with children <5 years (32.5 per 100,000 versus 25.4 per 100,000). Among Ontario’s public health regions, the hospitalization incidence varied from 19.3 per 100,000 to 36.1 per 100,000.

Age-stratified trends consistently aligned with overall age-standardized trends in, material resources, housing and dwellings, and age and labour force indices (**Figure S3**). Specifically, among those aged ≥65, the hospitalization rate increased twofold from increasing households and dwellings-related marginalization (37.3 per 100,000 versus 76.7 per 100,000). This twofold increase was also observed between the areas with the lowest and highest concentrations of the racialized and immigrant populations index. However, for those <5 years old, a decrease in hospitalization rates was observed with increasing neighbourhood-level quintile of racialized and immigrant populations marginalization (376.3 per 100,000 versus 240.7 per 100,000). RSV hospitalization rates also increased with increasing households and dwellings-related marginalization (251.9 per 100,000 versus 302.4 per 100,000). These gradients were also mirrored consistently across age groups.

### In-hospital outcomes

Among all RSV-related hospital admissions, 15.1% were admitted to the ICU and 3.5% experienced an in-hospital death (**Table 3**). ICU admissions were most common among those between the age 18-49 years old (27.6%). Death in hospital was more common among those aged ≥65 (7.0%) and the median length of stay for RSV-related hospitalization was longest in this age group as well, followed by the 50-64 age group (**Table S1**). Among patients aged ≥65 who died in hospital, the median length of stay was longest for those aged 75-84 years, followed by 65-74 years old (**Table S2**).

**Table 3:**
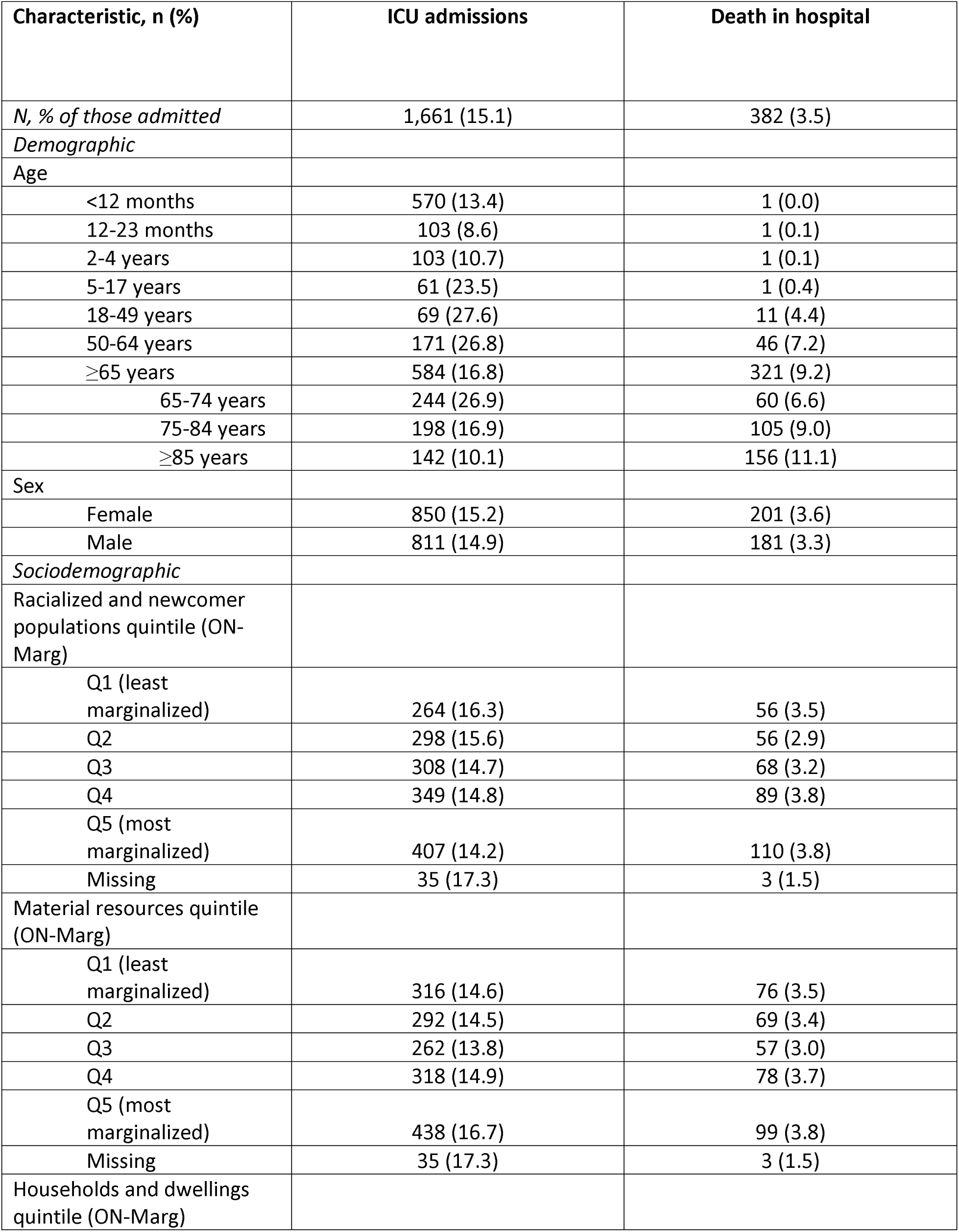
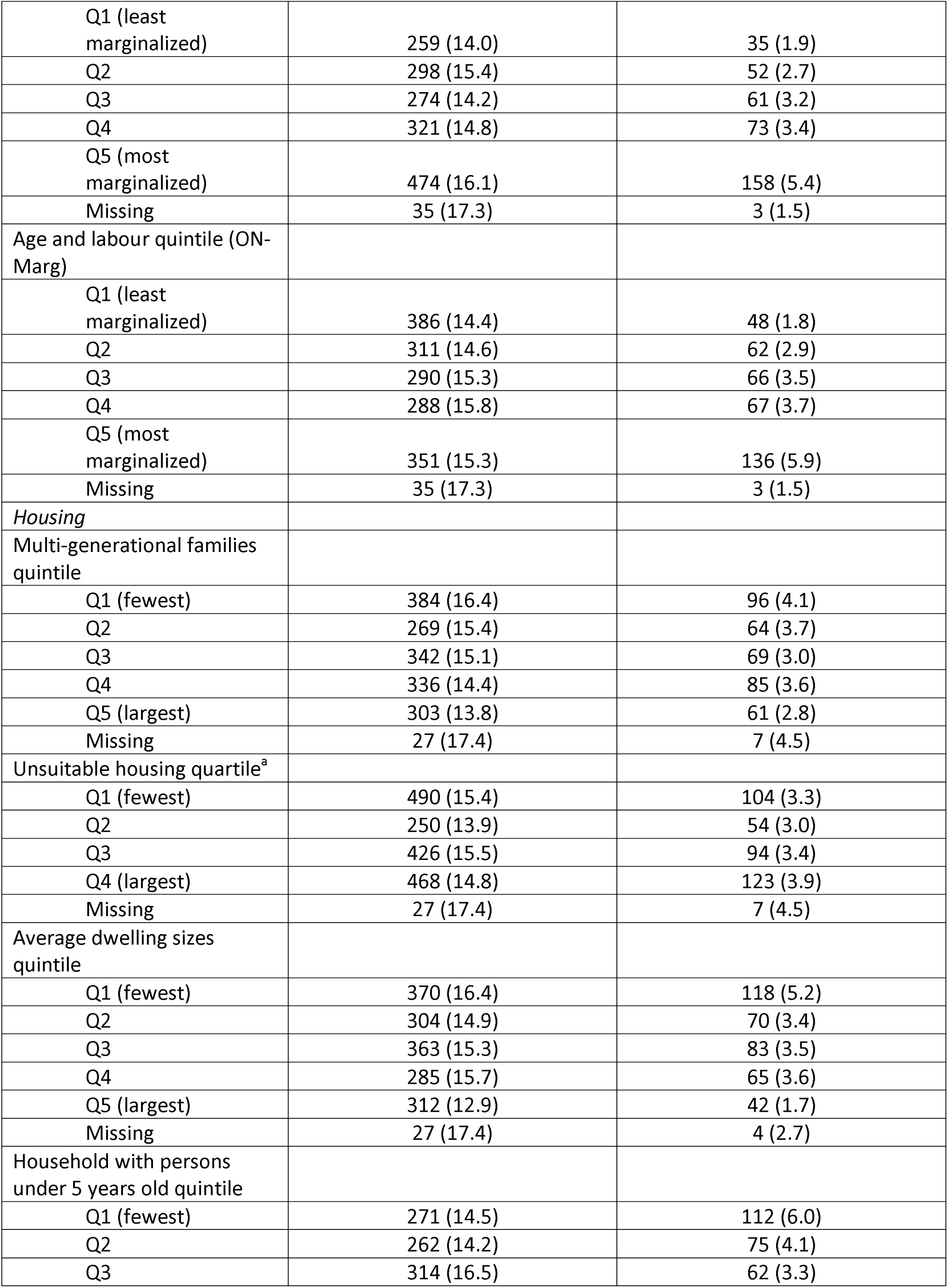
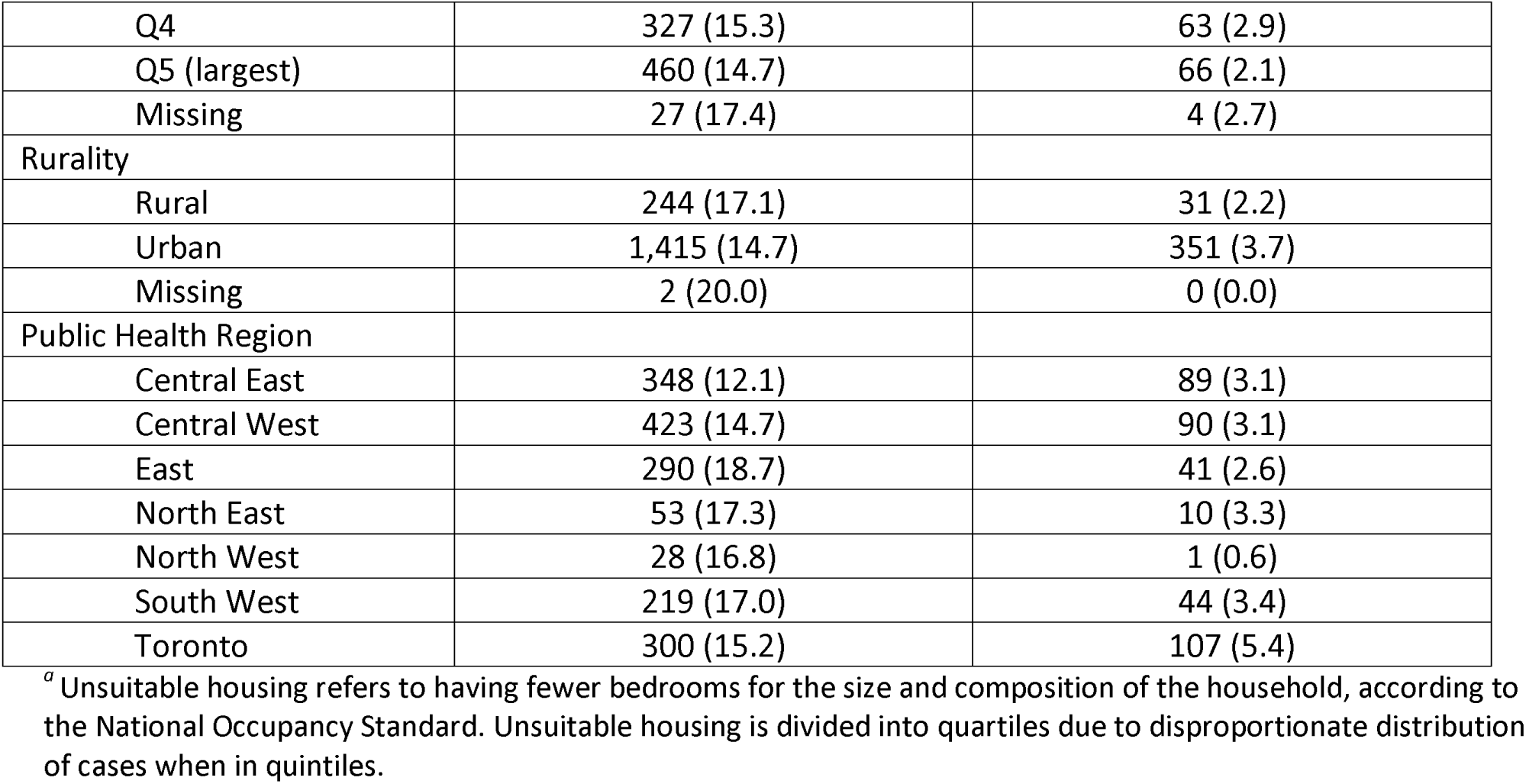
In-hospital outcomes following RSV-related hospitalization from September 1, 2016 to August 31, 2019.

| Characteristic, n (%) | ICU admissions | Death in hospital |
| --- | --- | --- |
| *N, % of those admitted* | 1,661 (15.1) | 382 (3.5) |
| *Demographic* |  |  |
| Age |  |  |
| <12 months | 570 (13.4) | 1 (0.0) |
| 12-23 months | 103 (8.6) | 1 (0.1) |
| 2-4 years | 103 (10.7) | 1 (0.1) |
| 5-17 years | 61 (23.5) | 1 (0.4) |
| 18-49 years | 69 (27.6) | 11 (4.4) |
| 50-64 years | 171 (26.8) | 46 (7.2) |
| ≥65 years | 584 (16.8) | 321 (9.2) |
| 65-74 years | 244 (26.9) | 60 (6.6) |
| 75-84 years | 198 (16.9) | 105 (9.0) |
| ≥85 years | 142 (10.1) | 156 (11.1) |
| Sex |  |  |
| Female | 850 (15.2) | 201 (3.6) |
| Male | 811 (14.9) | 181 (3.3) |
| *Sociodemographic* |  |  |
| Racialized and newcomer populations quintile (ON-Marg) |  |  |
| Q1 (least marginalized) | 264 (16.3) | 56 (3.5) |
| Q2 | 298 (15.6) | 56 (2.9) |
| Q3 | 308 (14.7) | 68 (3.2) |
| Q4 | 349 (14.8) | 89 (3.8) |
| Q5 (most marginalized) | 407 (14.2) | 110 (3.8) |
| Missing | 35 (17.3) | 3 (1.5) |
| Material resources quintile (ON-Marg) |  |  |
| Q1 (least marginalized) | 316 (14.6) | 76 (3.5) |
| Q2 | 292 (14.5) | 69 (3.4) |
| Q3 | 262 (13.8) | 57 (3.0) |
| Q4 | 318 (14.9) | 78 (3.7) |
| Q5 (most marginalized) | 438 (16.7) | 99 (3.8) |
| Missing | 35 (17.3) | 3 (1.5) |
| Households and dwellings quintile (ON-Marg) |  |  |
| Q1 (least marginalized) | 259 (14.0) | 35 (1.9) |
| Q2 | 298 (15.4) | 52 (2.7) |
| Q3 | 274 (14.2) | 61 (3.2) |
| Q4 | 321 (14.8) | 73 (3.4) |
| Q5 (most marginalized) | 474 (16.1) | 158 (5.4) |
| Missing | 35 (17.3) | 3 (1.5) |
| Age and labour quintile (ON-Marg) |  |  |
| Q1 (least marginalized) | 386 (14.4) | 48 (1.8) |
| Q2 | 311 (14.6) | 62 (2.9) |
| Q3 | 290 (15.3) | 66 (3.5) |
| Q4 | 288 (15.8) | 67 (3.7) |
| Q5 (most marginalized) | 351 (15.3) | 136 (5.9) |
| Missing | 35 (17.3) | 3 (1.5) |
| *Housing* |  |  |
| Multi-generational families quintile |  |  |
| Q1 (fewest) | 384 (16.4) | 96 (4.1) |
| Q2 | 269 (15.4) | 64 (3.7) |
| Q3 | 342 (15.1) | 69 (3.0) |
| Q4 | 336 (14.4) | 85 (3.6) |
| Q5 (largest) | 303 (13.8) | 61 (2.8) |
| Missing | 27 (17.4) | 7 (4.5) |
| Unsuitable housing quartile<sup>a</sup> |  |  |
| Q1 (fewest) | 490 (15.4) | 104 (3.3) |
| Q2 | 250 (13.9) | 54 (3.0) |
| Q3 | 426 (15.5) | 94 (3.4) |
| Q4 (largest) | 468 (14.8) | 123 (3.9) |
| Missing | 27 (17.4) | 7 (4.5) |
| Average dwelling sizes quintile |  |  |
| Q1 (fewest) | 370 (16.4) | 118 (5.2) |
| Q2 | 304 (14.9) | 70 (3.4) |
| Q3 | 363 (15.3) | 83 (3.5) |
| Q4 | 285 (15.7) | 65 (3.6) |
| Q5 (largest) | 312 (12.9) | 42 (1.7) |
| Missing | 27 (17.4) | 4 (2.7) |
| Household with persons under 5 years old quintile |  |  |
| Q1 (fewest) | 271 (14.5) | 112 (6.0) |
| Q2 | 262 (14.2) | 75 (4.1) |
| Q3 | 314 (16.5) | 62 (3.3) |
| Q4 | 327 (15.3) | 63 (2.9) |
| Q5 (largest) | 460 (14.7) | 66 (2.1) |
| Missing | 27 (17.4) | 4 (2.7) |
| Rurality |  |  |
| Rural | 244 (17.1) | 31 (2.2) |
| Urban | 1,415 (14.7) | 351 (3.7) |
| Missing | 2 (20.0) | 0 (0.0) |
| Public Health Region |  |  |
| Central East | 348 (12.1) | 89 (3.1) |
| Central West | 423 (14.7) | 90 (3.1) |
| East | 290 (18.7) | 41 (2.6) |
| North East | 53 (17.3) | 10 (3.3) |
| North West | 28 (16.8) | 1 (0.6) |
| South West | 219 (17.0) | 44 (3.4) |
| Toronto | 300 (15.2) | 107 (5.4) |

<sup>a</sup> Unsuitable housing refers to having fewer bedrooms for the size and composition of the household, according to the National Occupancy Standard. Unsuitable housing is divided into quartiles due to disproportionate distribution of cases when in quintiles.

### Trends by RSV diagnosis

Among the 11,039 RSV-related hospitalizations, 6,755 (61.2%) had one of the four RSV ICD-10-CA codes as the most responsible diagnosis (**Table S3**). For those with RSV as their most responsible diagnosis, hospitalizations were also greatest among those aged <12 months (55.7%), ≥65 years (17.2%), and 12-23 months (13.0%). Cases most frequently occurred during the winter months: December (23.7%), January (29.2%), and February (17.9%). Trends across demographic characteristics and quintiles for sociodemographic and housing characteristics were consistent for RSV as the most responsible diagnosis and when considered in all diagnostic fields (**Table S4A and Table S4B**). A larger gradient from low to high quintiles of marginalization was observed for multi-generational families (15.2 per 100,000 versus 20.4 per 100,000) for RSV as the most responsible diagnosis; however, an opposite gradient was noted when looking at all diagnosis.

For those with RSV as the most responsible diagnosis, 11.3% were admitted to the ICU and 1.4% experienced in-hospital mortality (**Table S5**). Median length of stay was also highest among individuals ≥65 years with RSV as the most responsible diagnosis (**Table S1**) and not the most responsible diagnosis (**Table S6**). For cases where RSV was not the most responsible diagnosis, acute upper respiratory infection emerged as the most common diagnosis across all age groups, and individuals aged ≥65 also showed congestive heart failure among their top five most responsible diagnoses (**Table S7**).

## Discussion

Between 2016 to 2019, we examined the age distribution of RSV-related hospitalization by calendar month, with the highest incidence among those <12 months followed by 12-23 months, 2-4 years, and ≥65 years in January. We found higher rates of RSV-related hospitalization in neighbourhoods characterized by marginalization related to both material resources and households and dwellings-related, fewer households with children <5 years, and households with smaller dwelling sizes. These findings highlight how neighbourhood conditions may be associated with RSV-related hospitalization inequities.

Overall, RSV-related hospitalization rates were consistent with Canadian literature; however, the magnitude of RSV-related hospitalization rates appears lower than US estimates [18,29–31]. Direct comparisons of absolute rates are challenging given differences in study population, methodology, and time period. Trends in RSV-related hospitalizations varied by sociodemographic indices in our study neighbourhoods with higher marginalization of material resources exhibited higher RSV-related hospitalization rates than less marginalized areas. This observation aligns with research exploring the association between income, poverty, and RSV infection [32,33]. Lower parental education, an element within the material and resource index in our study, has also been associated with higher RSV infection [34]. This is consistent with findings from Fitzpatrick et al. (2021) which also found inequities in RSV-related hospitalizations among Ontario children across ON-Marg indices, noting that children born to recent immigrants had a lower risk of RSV-related hospitalization compared to Canadian-born populations. This phenomenon might be linked to the healthy immigrant effect or Canada’s selective entry criteria for healthy and highly educated immigrants [14,35]. Alternatively, limited access to care for racialized and newcomer populations may be due to systemic barriers or distrust, leading to a reluctance to seek medical attention for mild to moderate conditions. We also found that higher rates of RSV-related hospitalization in neighborhoods experiencing greater marginalization related to households and dwellings, including poor housing conditions, smaller dwellings, and housing crowding as noted by previous studies [14,34,36].

We also found that neighbourhoods with fewer households with children <5 years had higher hospitalization rates. A possible explanation is that first-time mothers in households without other children <5 years may not have been exposed to infectious diseases as extensively as multiparous women, leading to potentially lower transfer of protective antibodies to their first child during childbirth [37]. Families with 2 or more siblings had no association with sibling infections occurring in one or multiple siblings [38]. Additionally, neighbourhoods with higher proportions of multi-generational families and larger dwelling sizes had lower rates of RSV-related hospitalizations, which could be attributed to prior RSV exposures and the subsequent development of immunity within these larger family dynamics. This may also reflect the differential use of childcare where families with more older children and relatives use less childcare, thereby limiting a risk factor for childhood infection [38].

In our study, the proportion of deaths in-hospital was highest for older populations, which is consistent with other studies [31,39–41]. Further, the proportion of hospitalized cases admitted to the ICU was highest in those aged ≥85 years. This is likely due to RSV-hospitalized individuals in this age group likely being immunocompromised and potentially having more underlying high-risk conditions, such as chronic obstructive pulmonary disease (COPD), asthma, and congestive heart failure, which can heighten susceptibility to severe RSV infections [39]. In addition, ICU admission was highest among 18-49 year olds, which was unexpected since RSV-related hospitalizations are less common in younger adult populations. While RSV reinfection is common throughout adult life, healthy young adults produce mild symptoms following infection [42]. The higher rates of ICU admission among individuals aged 18-49 years could be attributed to comorbidities [43].

The study showcases notable strengths, foremost among them being its novel exploration of social risk factors in RSV-related hospitalization across all age groups, which is a unique contribution to our understanding of RSV in Canada. Unlike existing research that often focuses on limited age ranges, this study explores the entirety of the age spectrum, making its findings more comprehensive and applicable to diverse populations. Additionally, we examined a wide array of demographic variables, allowing for a nuanced understanding of the intricate interplay between social determinants and RSV-related hospitalization rates. We further expanded sociodemographic exposures to include an aggregate of sociodemographic factors using the ON-Marg index and more information on housing characteristics, shedding light on the interaction of social and environmental factors in disease outcomes. This study uses a population-based approach, which addresses the limitations seen in other studies, such as Medicaid-focused research in the United States, with limited generalizability across a spectrum of socioeconomic status. Findings from a population-based study may have broader implications in informing targeted interventions for a diverse range of socioeconomic groups. We also looked at both RSV diagnoses as all possible diagnoses and the most responsible diagnosis in our study to capture the full burden of RSV-related hospitalization. Relying solely on the most responsible diagnosis may not fully capture the true burden of RSV-related hospitalization, as cases might be coded for comorbidities like COPD exacerbation despite being primarily RSV-related [19]. In addition, relying on a validated algorithm to define RSV-related hospitalization can capture a wider range of cases, given the low rate of lab confirmation [19].

However, this study is also subject to some limitations. First, our study focuses solely on hospitalized RSV cases, thus not providing a comprehensive assessment of the overall RSV burden. There may also be an underestimation of RSV hospital admissions since not all cases are tested or recorded as RSV. Our study may particularly underestimate RSV-related hospitalization rates in adults compared to pediatric age groups due to differences in algorithm sensitivity [44]. Further, focusing solely on hospitalized cases neglects the prevalence of outpatient RSV cases, omitting crucial information regarding ambulatory care that often precedes hospitalization and varies based on socioeconomic contexts. Although our study relies on ICD-10-coded outcomes, there is a possibility of misclassification; however, the RSV codes included have been previously validated and have high sensitivity and specificity [19]. Comorbidities were not examined in our study, which may contribute to differential rates across age and social risk factors. When interpreting area-level socioeconomic status, this is subjected to ecological fallacy which is a cautionary factor rather than a true limitation [45]. Associations observed at the aggregate level may not necessarily hold true at the individual level, emphasizing the need for careful interpretation of results.

## Conclusion

In conclusion, the highest incidence of RSV-related hospitalization was seen in neighbourhoods with greater material resources as well as households and dwellings-related marginalization. The highest burden of RSV was seen in those <5 and ≥65 years of age. Improving our understanding of the relative burdens of RSV across the age continuum will aid the implementation of prevention efforts, particularly with the current expansions of existing vaccination recommendations and the development of new vaccines. Sociodemographic factors play a critical role in shaping RSV-related hospitalization, and these relationships can provide a baseline against which to measure the impact of future intervention efforts on health disparities and complement ongoing work that is being done with RSV burden. With upcoming vaccine decisions, baseline RSV-related hospitalization rates across sociodemographic factors can support the prioritization of high-risk groups and equity-informed decisions for future vaccine rollout and monitoring.

## Data Availability

Public Health Ontario cannot disclose underlying data. Information on the data request process is available at https://www.publichealthontario.ca/en/data-and-analysis/using-data/data-requests

