## Supplementary file for "Neighbourhood-level burden of social risk factors on respiratory syncytial virus hospitalization in Ontario, Canada, 2016-2019"

**Supplementary Material**

**Table S1**: Median length of stay of RSV-related hospitalization by age, stratified by diagnosis.

| **Age** | **All diagnosis of RSV** | | **Most responsible diagnosis of RSV** | |
| --- | --- | --- | --- | --- |
|  | **Median length of stay (days, IQR)** | **Median length of stay in ICU (days, IQR)** | **Median length of stay (days, IQR)** | **Median length of stay in ICU (days, IQR)** |
| <12 months | 3.0 (2.0-5.0) | 4.0 (2.0-6.0) | 3.0 (2.0-4.0) | 3.0 (2.0-5.0) |
| 12-23 months | 2.0 (2.0-4.0) | 3.0 (2.0-5.0) | 2.0 (2.0-4.0) | 3.0 (2.0-5.0) |
| 2-4 years | 3.0 (2.0-4.0) | 3.0 (2.0-6.0) | 3.0 (2.0-4.0) | 3.0 (2.0-6.0) |
| 5-17 years | 4.0 (2.0-7.0) | 3.0 (1.0-8.0) | 3.0 (2.0-6.0) | 4.0 (2.0-9.0) |
| 18-49 years | 5.5 (3.0-12.0) | 4.0 (2.0-9.0) | 5.0 (3.0-9.0) | 4.0 (2.0-9.0) |
| 50-64 years | 6.0 (3.0-13.0) | 6.0 (3.0-10.0) | 5.0 (2.0-8.0) | 4.5 (1.0-7.0) |
| ≥65 years | 7.0 (4.0-13.0) | 4.0 (2.0-7.0) | 6.0 (3.0-10.0) | 4.0 (2.0-7.0) |
| 65-74 years | 6.0 (4.0-13.0) | 5.0 (2.0-8.0) | 6.0 (3.0-9.0) | 4.0 (2.0-7.0) |
| 75-84 years | 7.0 (4.0-12.0) | 4.0 (2.0-7.0) | 6.0 (3.0-10.0) | 4.0 (2.0-7.0) |
| ≥85 years | 7.0 (4.0-13.0) | 4.0 (2.0-7.0) | 6.0 (4.0-11.0) | 3.0 (2.0-6.0) |

**Table S2**: Median length of stay for patients aged 65+ years who died in hospital by diagnosis

| **Age** | **All diagnosis of RSV, n=321** | | | **Most responsible diagnosis of RSV, n=81** | | |
| --- | --- | --- | --- | --- | --- | --- |
|  | **Median length of stay (days, IQR)** | **Median length of stay in ICU (days, IQR)** | **% ICU admission,**  ***n (%)*** | **Median length of stay (days, IQR)** | **Median length of stay in ICU (days, IQR)** | **% ICU admission,**  ***n (%)*** |
| 65-74 years | 10.0 (4.5-19.0) | 5.0 (2.0-8.0) | 35 (58.3) | 7.0 (4.0-8.0) | 2.0 (1.5-10.5) | 8 (61.5) |
| 75-84 years | 13.0 (6.0-28.0) | 5.0 (2.0-9.0) | 51 (48.6) | 7.0 (4.0-17.0) | 6.0 (4.0-9.0) | 9 (39.1) |
| ≥85 years | 7.0 (3.5-16.5) | 4.0 (1.0-8.0) | 37 (23.7) | 7.0 (4.0-13.0) | 2.5 (1.0-10.0) | 6 (13.3) |

**Table S3**: Characteristics of patients hospitalized with RSV as the most responsible diagnosis in Ontario from September 1, 2016 to August 31, 2019.

| **Characteristics, n (%)** | **Individuals admitted with RSV as most responsible diagnosis (N = 6,755)** |
| --- | --- |
| *Demographic* |  |
| Age |  |
| <12 months | 3,760 (55.7) |
| 12-23 months | 880 (13.0) |
| 2-4 years | 576 (8.5) |
| 5-17 years | 83 (1.2) |
| 18-49 years | 97 (1.4) |
| 50-64 years | 198 (2.9) |
| ≥65 years | 1,161 (17.2) |
| 65-74 years | 254 (3.8) |
| 75-84 years | 373 (5.5) |
| ≥85 years | 534 (7.9) |
| Sex |  |
| Female | 3,238 (47.9) |
| Male | 3,517 (52.1) |
| Season |  |
| 2016-17 | 2,119 (31.4) |
| 2017-18 | 2,061 (30.5) |
| 2018-19 | 2,575 (38.1) |
| Month |  |
| September | 33 (0.5) |
| October | 100 (1.5) |
| November | 567 (8.4) |
| December | 1,842 (27.3) |
| January | 1,971 (29.2) |
| February | 1,209 (17.9) |
| March | 618 (9.2) |
| April | 287 (4.3) |
| May | 80 (1.2) |
| June | 21 (0.3) |
| July | 15 (0.2) |
| August | 12 (0.2) |
| *Sociodemographic* |  |
| Racialized and newcomer populations quintile (ON-Marg) |  |
| Q1 (least marginalized) | 991 (14.7) |
| Q2 | 1,153 (17.1) |
| Q3 | 1,240 (18.4) |
| Q4 | 1,471 (21.8) |
| Q5 (most marginalized) | 1,752 (25.9) |
| Missing | 148 (2.2) |
| Material resources quintile (ON-Marg) |  |
| Q1 (least marginalized) | 1,343 (19.9) |
| Q2 | 1,223 (18.1) |
| Q3 | 1,174 (17.4) |
| Q4 | 1,290 (19.1) |
| Q5 (most marginalized) | 1,577 (23.4) |
| Missing | 148 (2.2) |
| Households and dwellings quintile (ON-Marg) |  |
| Q1 (least marginalized) | 1,240 (18.4) |
| Q2 | 1,266 (18.7) |
| Q3 | 1,212 (17.9) |
| Q4 | 1,299 (19.2) |
| Q5 (most marginalized) | 1,590 (23.5) |
| Missing | 148 (2.2) |
| Age and labour quintile (ON-Marg) |  |
| Q1 (least marginalized) | 1,790 (26.5) |
| Q2 | 1,329 (19.7) |
| Q3 | 1,166 (17.3) |
| Q4 | 1,089 (16.1) |
| Q5 (most marginalized) | 1,233 (18.3) |
| Missing | 148 (2.2) |
| *Housing* |  |
| Multi-generational families quintile |  |
| Q1 (fewest) | 1,366 (20.2) |
| Q2 | 1,041 (15.4) |
| Q3 | 1,387 (20.5) |
| Q4 | 1,449 (21.5) |
| Q5 (largest) | 1,406 (20.8) |
| Missing | 106 (1.6) |
| Unsuitable housing quartile^a^ |  |
| Q1 (fewest) | 1,959 (29.0) |
| Q2 | 1,087 (16.1) |
| Q3 | 1,675 (24.8) |
| Q4 (largest) | 1,928 (28.5) |
| Missing | 106 (1.6) |
| Average dwelling sizes quintile |  |
| Q1 (fewest) | 1,191 (17.6) |
| Q2 | 1,234 (18.3) |
| Q3 | 1,476 (21.9) |
| Q4 | 1,123 (16.6) |
| Q5 (largest) | 1,626 (24.1) |
| Missing | 105 (1.6) |
| Household with persons under 5 years old quintile |  |
| Q1 (fewest) | 980 (14.5) |
| Q2 | 1,064 (15.8) |
| Q3 | 1,168 (17.3) |
| Q4 | 1,360 (20.1) |
| Q5 (largest) | 2,078 (30.8) |
| Missing | 105 (1.6) |
| Rurality |  |
| Rural | 962 (14.2) |
| Urban | 5,790 (85.7) |
| Missing | 3 (0.0) |
| Public Health Region |  |
| Central East | 1,863 (27.6) |
| Central West | 1,779 (26.3) |
| East | 881 (13.0) |
| North East | 203 (3.0) |
| North West | 130 (1.9) |
| South West | 792 (11.7) |
| Toronto | 1,107 (16.4) |

*^a^* Unsuitable housing refers to having fewer bedrooms for the size and composition of the household, according to the National Occupancy Standard. Unsuitable housing is divided into quartiles due to disproportionate distribution of cases when in quintiles.

**Table S4A**: Crude annualized RSV-related hospitalization rates per 100,000 in Ontario by demographic characteristics with RSV as most responsible diagnostic from September 1, 2016 to August 31, 2019.

| **Characteristic** | **RSV-related hospitalizations, 2016-2019, n (%)** | **Ontario population,**  **2016** | **Annualized RSV-related hospitalization rate per 100,000**^a^ **(95% CI)** |
| --- | --- | --- | --- |
| *Demographic* |  |  |  |
| Age |  |  |  |
| <12 months | 3,760 (55.7) | 135,080 | 927.8 (898.2, 957.5) |
| 12-23 months | 880 (13.0) | 135,875 | 215.9 (201.6, 230.1) |
| 2-4 years | 576 (8.5) | 426,510 | 45.0 (41.3, 48.7) |
| 5-17 years | 83 (1.2) | 1,985,175 | 1.4 (1.1, 1.7) |
| 18-49 years | 97 (1.4) | 5,618,050 | 0.6 (0.5, 0.7) |
| 50-64 years | 198 (2.9) | 2,892,625 | 2.3 (2.0, 2.6) |
| ≥65 years | 1,161 (17.2) | 2,249,625 | 17.2 (16.2, 18.2) |
| 65-74 years | 254 (3.8) | 1,265,375 | 6.7 (5.9, 7.5) |
| 75-84 years | 373 (5.5) | 683,330 | 18.2 (16.3, 20.0) |
| ≥85 years | 534 (7.9) | 300,920 | 59.2 (54.1, 64.2) |
| Sex |  |  |  |
| Female | 3,238 (47.9) | 6,888,235 | 15.7 (15.1, 16.2) |
| Male | 3,517 (52.1) | 6,554,705 | 17.9 (17.3, 18.5) |
| Season^b^ |  |  |  |
| 2016-17 | 2,119 (31.4) | 13,448,155 | 15.8 (15.1, 16.4) |
| 2017-18 | 2,061 (30.5) | 13,448,155 | 15.3 (14.7, 16.0) |
| 2018-19 | 2,575 (38.1) | 13,448,155 | 19.1 (18.4, 19.9) |
| Month |  |  |  |
| September | 33 (0.5) | 13,448,155 | 0.1 (0.1, 0.1) |
| October | 100 (1.5) | 13,448,155 | 0.2 (0.2, 0.3) |
| November | 567 (8.4) | 13,448,155 | 1.4 (1.3, 1.5) |
| December | 1,842 (27.3) | 13,448,155 | 4.6 (4.4, 4.8) |
| January | 1,971 (29.2) | 13,448,155 | 4.9 (4.7, 5.1) |
| February | 1,209 (17.9) | 13,448,155 | 3.0 (2.8, 3.2) |
| March | 618 (9.2) | 13,448,155 | 1.5 (1.4, 1.7) |
| April | 287 (4.3) | 13,448,155 | 0.7 (0.6, 0.8) |
| May | 80 (1.2) | 13,448,155 | 0.2 (0.2, 0.2) |
| June | 21 (0.3) | 13,448,155 | 0.1 (0.0, 0.1) |
| July | 15 (0.2) | 13,448,155 | 0.0 (0.0, 0.1) |
| August | 12 (0.2) | 13,448,155 | 0.0 (0.0, 0.0) |

**Missing rates were not included in table.*

^a^ Annualized rate is calculated using the cumulative number of RSV-related hospitalizations from September 1, 2016 to August 31, 2019 as the numerator and the 2016 Ontario Census multiplied by 3 as the denominator.

^b^ Rate for season is calculated using the cumulative number of RSV-related hospitalizations from September 1, 2016 to August 31, 2019 as the numerator and the 2016 Ontario Census as the denominator.

**Table S4B**: Age-standardized annualized RSV-related hospitalization rates per 100,000 in Ontario by sociodemographic and housing characteristics with RSV as most responsible diagnostic from September 1, 2016 to August 31, 2019.

| **Characteristic** | **RSV-related hospitalizations, 2016-2019,**  **n (%)** | **Ontario population, 2016** | **Age-standardized annualized RSV-related hospitalization rate per 100,000**^a^ **(95% CI)** | **Rate ratio**  **(95% CI)** |
| --- | --- | --- | --- | --- |
| *Overall* | 6,755 | 13,448,155 | 17.2 (16.6, 17.9) | - |
| *Sociodemographic* |  |  |  |  |
| Racialized and newcomer populations quintile (ON-Marg) |  |  |  |  |
| Q1 (least marginalized) | 991 (14.7) | 2,074,090 | 19.5 (17.8, 21.2) | 1.0 (reference) |
| Q2 | 1,153 (17.1) | 2,209,735 | 19.1 (17.5, 20.7) | 1.0 (1.0, 1.0) |
| Q3 | 1,240 (18.4) | 2,392,660 | 18.1 (16.6, 19.5) | 0.9 (0.9, 0.9) |
| Q4 | 1,471 (21.8) | 2,827,935 | 18.0 (16.5, 19.4) | 0.9 (0.9, 0.9) |
| Q5 (most marginalized) | 1,752 (25.9) | 2,074,090 | 14.7 (13.6, 15.8) | 0.8 (0.8, 0.7) |
| Material resources quintile (ON-Marg) |  |  |  |  |
| Q1 (least marginalized) | 1,343 (19.9) | 3,027,125 | 15.2 (14.0, 16.5) | 1.0 (reference) |
| Q2 | 1,223 (18.1) | 2,832,470 | 15.0 (13.8, 16.3) | 1.0 (1.0, 1.0) |
| Q3 | 1,174 (17.4) | 2,546,320 | 16.5 (15.1, 17.8) | 1.1 (1.1, 1.1) |
| Q4 | 1,290 (19.1) | 2,432,020 | 18.8 (17.3, 20.3) | 1.2 (1.2, 1.2) |
| Q5 (most marginalized) | 1,577 (23.4) | 2,540,425 | 19.8 (18.3, 21.3) | 1.3 (1.3, 1.3) |
| Households and dwellings quintile (ON-Marg) |  |  |  |  |
| Q1 (least marginalized) | 1,240 (18.4) | 2,968,800 | 14.1 (12.9, 15.3) | 1.0 (reference) |
| Q2 | 1,266 (18.7) | 2,534,625 | 17.2 (15.8, 18.6) | 1.2 (1.2, 1.2) |
| Q3 | 1,212 (17.9) | 2,392,230 | 17.3 (15.9, 18.7) | 1.2 (1.2, 1.2) |
| Q4 | 1,299 (19.2) | 2,405,570 | 18.2 (16.7, 19.6) | 1.3 (1.3, 1.3) |
| Q5 (most marginalized) | 1,590 (23.5) | 3,077,135 | 18.3 (16.9, 19.6) | 1.3 (1.3, 1.3) |
| Age and labour quintile (ON-Marg) |  |  |  |  |
| Q1 (least marginalized) | 1,790 (26.5) | 3,656,515 | 15.2 (14.0, 16.3) | 1.0 (reference) |
| Q2 | 1,329 (19.7) | 2,745,855 | 16.6 (15.3, 18.0) | 1.1 (1.1, 1.1) |
| Q3 | 1,166 (17.3) | 2,356,490 | 17.8 (16.3, 19.4) | 1.2 (1.2, 1.2) |
| Q4 | 1,089 (16.1) | 2,242,400 | 17.9 (16.4, 19.5) | 1.2 (1.2, 1.2) |
| Q5 (most marginalized) | 1,233 (18.3) | 2,377,250 | 20.4 (18.7, 22.1) | 1.3 (1.3, 1.4) |
| *Housing* |  |  |  |  |
| Multi-generational families quintile |  |  |  |  |
| Q1 (fewest) | 1,366 (20.2) | 2,749,085 | 15.2 (14.0, 16.3) | 1.0 (reference) |
| Q2 | 1,041 (15.4) | 2,094,330 | 16.6 (15.3, 18.0) | 1.1 (1.1, 1.1) |
| Q3 | 1,387 (20.5) | 2,732,730 | 17.8 (16.3, 19.4) | 1.2 (1.2, 1.2) |
| Q4 | 1,449 (21.5) | 2,840,820 | 17.9 (16.4, 19.5) | 1.2 (1.2, 1.2) |
| Q5 (largest) | 1,406 (20.8) | 3,021,505 | 20.4 (18.7, 22.1) | 1.3 (1.3, 1.4) |
| Unsuitable housing quartile^b^ |  |  |  |  |
| Q1 (fewest) | 1,959 (29.0) | 4,103,625 | 17.7 (16.6, 18.8) | 1.0 (reference) |
| Q2 | 1,087 (16.1) | 2,188,520 | 16.2 (14.8, 17.7) | 0.9 (0.9, 0.9) |
| Q3 | 1,675 (24.8) | 3,258,440 | 17.7 (16.4, 19.0) | 1.0 (1.0, 1.0) |
| Q4 (largest) | 1,928 (28.5) | 3,887,885 | 16.5 (15.4, 17.7) | 0.9 (0.9, 0.9) |
| Average dwelling sizes quintile |  |  |  |  |
| Q1 (fewest) | 1,191 (17.6) | 2,477,800 | 19.3 (17.7, 20.9) | 1.0 (reference) |
| Q2 | 1,234 (18.3) | 2,353,065 | 18.4 (16.9, 19.9) | 1.0 (1.0, 1.0) |
| Q3 | 1,476 (21.9) | 2,776,080 | 18.0 (16.6, 19.4) | 0.9 (0.9, 0.9) |
| Q4 | 1,123 (16.6) | 2,263,490 | 16.5 (15.1, 18.0) | 0.9 (0.9, 0.9) |
| Q5 (largest) | 1,626 (24.1) | 3,569,875 | 14.7 (13.5, 15.8) | 0.8 (0.8, 0.8) |
| Household with persons under 5 years old quintile |  |  |  |  |
| Q1 (fewest) | 980 (14.5) | 2,365,265 | 21.9 (19.9, 23.9) | 1.0 (reference) |
| Q2 | 1,064 (15.8) | 2,439,210 | 18.3 (16.7, 19.9) | 0.8 (0.8, 0.8) |
| Q3 | 1,168 (17.3) | 2,494,680 | 17.3 (15.9, 18.8) | 0.8 (0.8, 0.8) |
| Q4 | 1,360 (20.1) | 2,675,160 | 16.6 (15.3, 18.0) | 0.8 (0.8, 0.8) |
| Q5 (largest) | 2,078 (30.8) | 3,465,995 | 15.3 (14.2, 16.3) | 0.7 (0.7, 0.7) |
| Rurality |  |  |  |  |
| Rural | 962 (14.2) | 1,848,105 | 18.3 (16.7, 19.9) | 1.0 (reference) |
| Urban | 5,790 (85.7) | 11,600,050 | 17.1 (16.5, 17.8) | 0.9 (0.9, 1.0) |
| Public Health Region |  |  |  |  |
| Central East | 1,863 (27.6) | 3,985,675 | 16.1 (15.1, 17.2) | - |
| Central West | 1,779 (26.3) | 2,602,740 | 22.9 (21.3, 24.4) | - |
| East | 881 (13.0) | 1,762,675 | 17.5 (15.8, 19.2) | - |
| North East | 203 (3.0) | 550,490 | 13.4 (10.9, 15.9) | - |
| North West | 130 (1.9) | 226,980 | 18.7 (14.2, 23.3) | - |
| South West | 792 (11.7) | 1,581,065 | 16.8 (15.2, 18.5) | - |
| Toronto | 1,107 (16.4) | 2,730,685 | 14.0 (12.7, 15.4) | - |

**Missing rates were not included in table.*

^a^ Annualized rate is calculated using the cumulative number of RSV-related hospitalizations from September 1, 2016 to August 31, 2019 as the numerator and the 2016 Ontario Census multiplied by 3 as the denominator.

^b^ Unsuitable housing refers to having fewer bedrooms for the size and composition of the household, according to the National Occupancy Standard. Unsuitable housing is divided into quartiles due to disproportionate distribution of cases when in quintiles.

**Table S5**: In-hospital outcomes following RSV-related hospitalization with RSV as most responsible diagnostic from September 1, 2016 to August 31, 2019.

| **Characteristics, n (%)** | **ICU admissions** | **Death in hospital** |
| --- | --- | --- |
| *N, % of those admitted* | 766 (11.3) | 97 (1.4) |
| *Demographic* |  |  |
| Age |  |  |
| <12 months | 454 (12.1) | 0 (0.0) |
| 12-23 months | 69 (7.8) | 0 (0.0) |
| 2-4 years | 47 (8.2) | 1 (0.2) |
| 5-17 years | 19 (22.9) | 0 (0.0) |
| 18-49 years | 19 (19.6) | 4 (4.1) |
| 50-64 years | 34 (17.2) | 11 (5.6) |
| ≥65 years | 124 (10.7) | 81 (7.0) |
| 65-74 years | 43 (16.9) | 13 (5.1) |
| 75-84 years | 42 (11.3) | 23 (6.2) |
| ≥85 years | 39 (7.3) | 45 (8.4) |
| Sex |  |  |
| Female | 369 (11.4) | 48 (1.5) |
| Male | 397 (11.3) | 49 (1.4) |
| *Sociodemographic* |  |  |
| Racialized and newcomer populations quintile (ON-Marg) |  |  |
| Q1 (least marginalized) | 118 (11.9) | 12 (1.2) |
| Q2 | 129 (11.2) | 13 (1.1) |
| Q3 | 135 (10.9) | 15 (1.2) |
| Q4 | 166 (11.3) | 28 (1.9) |
| Q5 (most marginalized) | 192 (11.0) | 29 (1.7) |
| Missing | 26 (17.6) | 0 (0.0) |
| Material resources quintile (ON-Marg) |  |  |
| Q1 (least marginalized) | 165 (12.3) | 23 (1.7) |
| Q2 | 129 (10.5) | 19 (1.6) |
| Q3 | 120 (10.2) | 14 (1.2) |
| Q4 | 146 (11.3) | 26 (2.0) |
| Q5 (most marginalized) | 180 (11.4) | 15 (1.0) |
| Missing | 26 (17.6) | 0 (0.0) |
| Households and dwellings quintile (ON-Marg) |  |  |
| Q1 (least marginalized) | 141 (11.4) | 8 (0.6) |
| Q2 | 142 (11.2) | 14 (1.1) |
| Q3 | 132 (10.9) | 17 (1.4) |
| Q4 | 132 (10.2) | 21 (1.6) |
| Q5 (most marginalized) | 193 (12.1) | 37 (2.3) |
| Missing | 26 (17.6) | 0 (0.0) |
| Age and labour quintile (ON-Marg) |  |  |
| Q1 (least marginalized) | 201 (11.2) | 11 (0.6) |
| Q2 | 158 (11.9) | 14 (1.1) |
| Q3 | 132 (11.3) | 13 (1.1) |
| Q4 | 122 (11.2) | 17 (1.6) |
| Q5 (most marginalized) | 127 (10.3) | 42 (3.4) |
| Missing | 26 (17.6) | 0 (0.0) |
| *Housing* |  |  |
| Multi-generational families quintile |  |  |
| Q1 (fewest) | 169 (12.4) | 29 (2.1) |
| Q2 | 122 (11.7) | 11 (1.1) |
| Q3 | 164 (11.8) | 19 (1.4) |
| Q4 | 156 (10.8) | 23 (1.6) |
| Q5 (largest) | 136 (9.7) | 15 (1.1) |
| Missing | 19 (17.9) | 0 (0.0) |
| Unsuitable housing quartile^a^ |  |  |
| Q1 (fewest) | 216 (11.0) | 32 (1.6) |
| Q2 | 118 (10.9) | 9 (0.8) |
| Q3 | 201 (12.0) | 21 (1.3) |
| Q4 (largest) | 212 (11.0) | 35 (1.8) |
| Missing | 19 (17.9) | 0 (0.0) |
| Average dwelling sizes quintile |  |  |
| Q1 (fewest) | 144 (12.1) | 25 (2.1) |
| Q2 | 143 (11.6) | 23 (1.9) |
| Q3 | 155 (10.5) | 18 (1.2) |
| Q4 | 135 (12.0) | 17 (1.5) |
| Q5 (largest) | 170 (10.5) | 14 (0.9) |
| Missing | 19 (18.1) | 0 (0.0) |
| Household with persons under 5 years old quintile |  |  |
| Q1 (fewest) | 105 (8.8) | 36 (3.0) |
| Q2 | 98 (7.9) | 23 (1.9) |
| Q3 | 135 (9.1) | 13 (0.9) |
| Q4 | 161 (14.3) | 11 (1.0) |
| Q5 (largest) | 248 (15.3) | 14 (0.9) |
| Missing | 19 (18.1) | 0 (0.0) |
| Rurality |  |  |
| Rural | 134 (13.9) | 7 (0.7) |
| Urban | 631 (10.9) | 90 (1.6) |
| Missing | 1 (33.3) | 0 (0.0) |
| Public Health Region |  |  |
| Central East | 164 (8.8) | 22 (1.2) |
| Central West | 196 (11.0) | 20 (1.1) |
| East | 130 (14.8) | 6 (0.7) |
| North East | 17 (8.4) | 1 (0.5) |
| North West | 19 (14.6) | 1 (0.8) |
| South West | 113 (14.3) | 9 (1.1) |
| Toronto | 127 (11.5) | 38 (3.4) |

^a^ Unsuitable housing refers to having more household members occupying the same dwelling unit to live comfortably together, according to the National Occupancy Standard. Unsuitable housing is divided into quartiles due to disproportionate distribution of cases when in quintiles.

**Table S6**: Hospital outcomes of RSV-related hospitalization for patients aged 65+ years without RSV as their most responsible diagnosis (n = 4284).

| **Age** | **Median length of stay**  **(days, IQR)** | **Median length of stay in ICU**  **(days, IQR)** | **% ICU admissions,**  ***n (%)*** | **% Death in hospital,**  ***n (%)*** |
| --- | --- | --- | --- | --- |
| 65-74 years | 7.0 (4.0-15.0) | 5.0 (2.0-8.0) | 201 (30.7) | 47 (7.2) |
| 75-84 years | 7.0 (4.0-14.0) | 4.0 (2.0-7.0) | 156 (19.5) | 82 (10.3) |
| ≥85 years | 8.0 (4.0-16.0) | 4.0 (2.0-7.0) | 103 (11.9) | 111 (12.8) |

**Table S7**: Top 5 most responsible diagnosis of patients without RSV as their most responsible diagnosis by age (n = 4284).

| **Age, Most responsible diagnosis, n (%)** | **Non-RSV most responsible diagnosis** |
| --- | --- |
| <5 years |  |
| (J069) ACUTE UPPER RESPIRATORY INFECTION, UNSPECIFIED | 166 (13.9) |
| (J4500) PREDOMINANTLY ALLERGIC ASTHMA WITHOUT STATED STATUS ASTHMATICUS | 156 (13.1) |
| (J159) BACTERIAL PNEUMONIA, UNSPECIFIED | 131 (11.0) |
| (J189) PNEUMONIA, UNSPECIFIED | 93 (7.8) |
| (J22) UNSPECIFIED ACUTE LOWER RESPIRATORY INFECTION | 50 (4.2) |
| 5-64 years |  |
| (J440) CHRONIC OBSTRUCTIVE PULMONARY DISEASE WITH ACUTE LOWER RESPIRATORY INFECTION | 107 (13.9) |
| (J441) CHRONIC OBSTRUCTIVE PULMONARY DISEASE WITH ACUTE EXACERBATION, UNSPECIFIED | 69 (9.0) |
| (J069) ACUTE UPPER RESPIRATORY INFECTION, UNSPECIFIED | 59 (7.7) |
| (J4590) ASTHMA, UNSPECIFIED, WITHOUT STATED STATUS ASTHMATICUS | 27 (3.5) |
| (J4500) PREDOMINANTLY ALLERGIC ASTHMA WITHOUT STATED STATUS ASTHMATICUS | 26 (3.4) |
| ≥65 years |  |
| (J440) CHRONIC OBSTRUCTIVE PULMONARY DISEASE WITH ACUTE LOWER RESPIRATORY INFECTION | 511 (22.0) |
| (J441) CHRONIC OBSTRUCTIVE PULMONARY DISEASE WITH ACUTE EXACERBATION, UNSPECIFIED | 316 (13.6) |
| (J069) ACUTE UPPER RESPIRATORY INFECTION, UNSPECIFIED | 212 (9.1) |
| (I500) CONGESTIVE HEART FAILURE | 159 (6.9) |
| (J4590) ASTHMA, UNSPECIFIED, WITHOUT STATED STATUS ASTHMATICUS | 57 (2.5) |

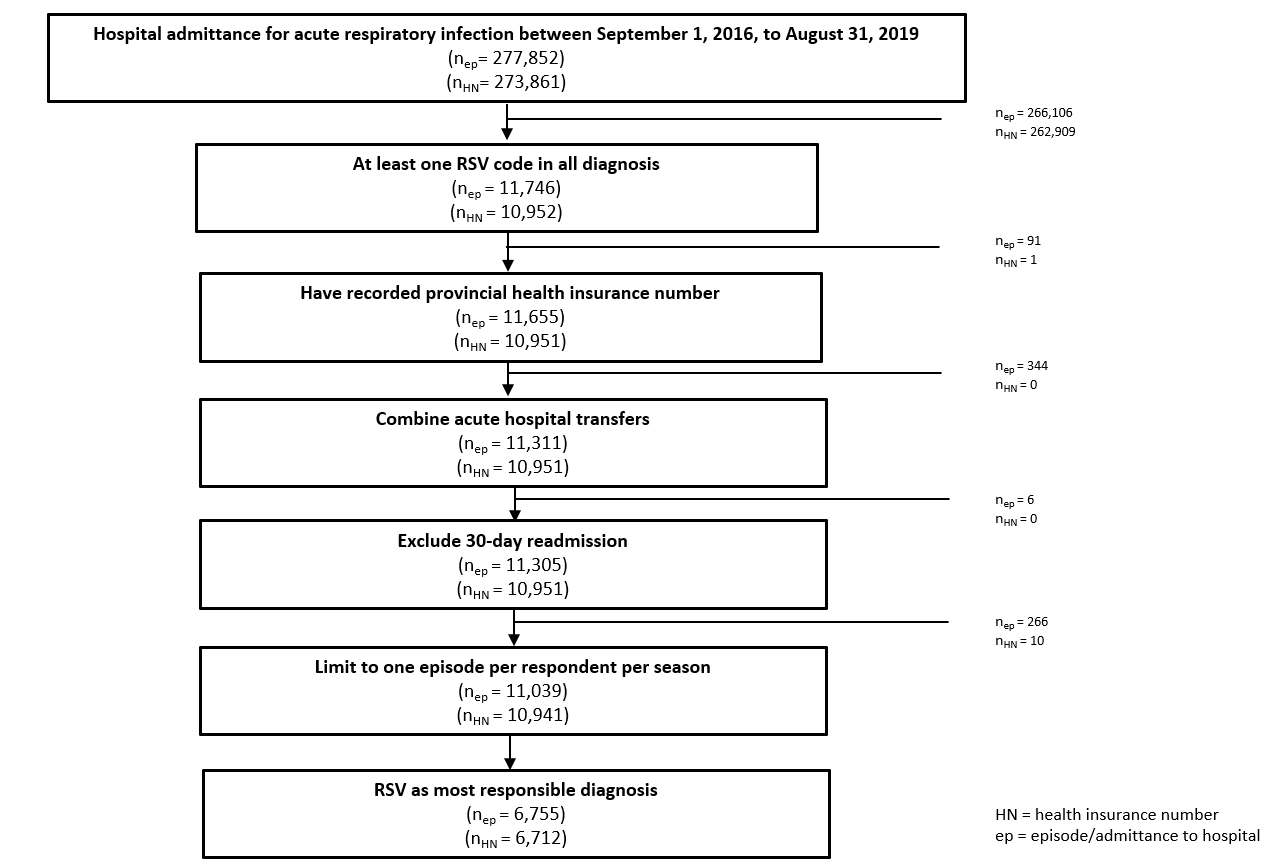

**Figure S1:** Cohort design flow chart.

Cumulative RSV hospitalization (%)

|  | <12 months | 12-23 months | 2-4 years | 5-17 years | 18-49 years | 50-64 years | ≥65 years |
| --- | --- | --- | --- | --- | --- | --- | --- |
| September | 0.5 | 0.4 | 0.6 | 0.4 | 0 | 0.6 | 0.5 |
| October | 2.3 | 2.4 | 3 | 2.3 | 1.6 | 1.4 | 1.4 |
| November | 10.8 | 12.8 | 17.6 | 11.1 | 4.4 | 5.6 | 5.9 |
| December | 37.4 | 46.5 | 54.9 | 42.6 | 27.2 | 24.9 | 24.8 |
| January | 67.5 | 72.2 | 78.2 | 68 | 56 | 59.1 | 59.5 |
| February | 85.3 | 88 | 92.6 | 85.3 | 73.6 | 78.1 | 78.8 |
| March | 93.8 | 95.8 | 96.6 | 92.2 | 86.4 | 91.3 | 92.4 |
| April | 97.8 | 98.6 | 98.1 | 96.8 | 96.4 | 96.2 | 98 |
| May | 99.1 | 99.4 | 99.5 | 98.7 | 98.4 | 98.2 | 99.3 |
| June | 99.5 | 99.8 | 99.8 | 99.5 | 98.4 | 99 | 99.6 |
| July | 99.7 | 100.1 | 99.8 | 99.5 | 98.8 | 99.5 | 99.8 |
| August | 99.9 | 100.2 | 100.1 | 99.9 | 100 | 100.1 | 100.1 |

**Figure S2:** Cumulative proportion of RSV-related hospitalization by month from September 1, 2016 to August 31, 2019, stratified by age.

<5 years

Incidence per 100,000

|  | Rate |
| --- | --- |
| Racialized and newcomer populations Q1 | 376.3 |
| Racialized and newcomer populations Q2 | 356 |
| Racialized and newcomer populations Q3 | 323 |
| Racialized and newcomer populations Q4 | 299.9 |
| Racialized and newcomer populations Q5 | 240.7 |
| Material resources Q1 | 262.1 |
| Material resources Q2 | 266 |
| Material resources Q3 | 295.4 |
| Material resources Q4 | 337.2 |
| Material resources Q5 | 357 |
| Households and dwellings Q1 | 251.9 |
| Households and dwellings Q2 | 317.9 |
| Households and dwellings Q3 | 319.4 |
| Households and dwellings Q4 | 333.2 |
| Households and dwellings Q5 | 302.4 |
| Age and labour Q1 | 266.6 |
| Age and labour Q2 | 291.9 |
| Age and labour Q3 | 319.3 |
| Age and labour Q4 | 329.1 |
| Age and labour Q5 | 363.7 |
| Multi-generational families Q1 | 337 |
| Multi-generational families Q2 | 314.6 |
| Multi-generational families Q3 | 310.7 |
| Multi-generational families Q4 | 299 |
| Multi-generational families Q5 | 263.9 |
| Unsuitable housing Q1 | 328.4 |
| Unsuitable housing Q2 | 293.6 |
| Unsuitable housing Q3 | 316.6 |
| Unsuitable housing Q4 | 272.3 |
| Average dwelling sizes Q1 | 341.7 |
| Average dwelling sizes Q2 | 344.6 |
| Average dwelling sizes Q3 | 323.9 |
| Average dwelling sizes Q4 | 293.7 |
| Average dwelling sizes Q5 | 251.8 |
| Household with persons under 5 years old Q1 | 391.3 |
| Household with persons under 5 years old Q2 | 329.9 |
| Household with persons under 5 years old Q3 | 307.1 |
| Household with persons under 5 years old Q4 | 299 |
| Household with persons under 5 years old Q5 | 272.9 |

|  |  |
| --- | --- |
| 376.3 | 0.7 |
| 356 | 1.8 |
| 323 | 3 |
| 299.9 | 4.2 |
| 240.7 | 5.4 |
| 262.1 | 6.6 |
| 266 | 7.8 |
| 295.4 | 9 |
| 337.2 | 10.2 |
| 357 | 11.4 |
| 251.9 | 12.6 |
| 317.9 | 13.8 |
| 319.4 | 15 |
| 333.2 | 16.2 |
| 302.4 | 17.4 |
| 266.6 | 18.6 |
| 291.9 | 19.8 |
| 319.3 | 21 |
| 329.1 | 22.2 |
| 363.7 | 23.4 |
| 337 | 24.6 |
| 314.6 | 25.8 |
| 310.7 | 27 |
| 299 | 28.2 |
| 263.9 | 29.4 |
| 328.4 | 30.6 |
| 293.6 | 31.8 |
| 316.6 | 33 |
| 272.3 | 34.2 |
| 341.7 | 35.4 |
| 344.6 | 36.6 |
| 323.9 | 37.8 |
| 293.7 | 39 |
| 251.8 | 40.2 |
| 391.3 | 41.4 |
| 329.9 | 42.6 |
| 307.1 | 43.8 |
| 299 | 45 |
| 272.9 | 46.2 |

5-64 years

Incidence per 100,000

|  | Rate |
| --- | --- |
| Racialized and newcomer populations Q1 | 3.4 |
| Racialized and newcomer populations Q2 | 3.8 |
| Racialized and newcomer populations Q3 | 3.7 |
| Racialized and newcomer populations Q4 | 3.4 |
| Racialized and newcomer populations Q5 | 3.7 |
| Material resources Q1 | 3.2 |
| Material resources Q2 | 2.9 |
| Material resources Q3 | 3 |
| Material resources Q4 | 4 |
| Material resources Q5 | 5.2 |
| Households and dwellings Q1 | 2.7 |
| Households and dwellings Q2 | 2.9 |
| Households and dwellings Q3 | 3.6 |
| Households and dwellings Q4 | 4.4 |
| Households and dwellings Q5 | 4.5 |
| Age and labour Q1 | 3.2 |
| Age and labour Q2 | 3.5 |
| Age and labour Q3 | 3.9 |
| Age and labour Q4 | 3.7 |
| Age and labour Q5 | 4.1 |
| Multi-generational families Q1 | 3.6 |
| Multi-generational families Q2 | 4 |
| Multi-generational families Q3 | 3.6 |
| Multi-generational families Q4 | 3.5 |
| Multi-generational families Q5 | 3.4 |
| Unsuitable housing Q1 | 3.5 |
| Unsuitable housing Q2 | 3.2 |
| Unsuitable housing Q3 | 3.7 |
| Unsuitable housing Q4 | 3.9 |
| Average dwelling sizes Q1 | 4.3 |
| Average dwelling sizes Q2 | 4.2 |
| Average dwelling sizes Q3 | 3.7 |
| Average dwelling sizes Q4 | 3.3 |
| Average dwelling sizes Q5 | 2.9 |
| Household with persons under 5 years old Q1 | 3.8 |
| Household with persons under 5 years old Q2 | 3.6 |
| Household with persons under 5 years old Q3 | 3.8 |
| Household with persons under 5 years old Q4 | 3.6 |
| Household with persons under 5 years old Q5 | 3.4 |

|  |  |
| --- | --- |
| 3.4 | 0.7 |
| 3.8 | 1.8 |
| 3.7 | 3 |
| 3.4 | 4.2 |
| 3.7 | 5.4 |
| 3.2 | 6.6 |
| 2.9 | 7.8 |
| 3 | 9 |
| 4 | 10.2 |
| 5.2 | 11.4 |
| 2.7 | 12.6 |
| 2.9 | 13.8 |
| 3.6 | 15 |
| 4.4 | 16.2 |
| 4.5 | 17.4 |
| 3.2 | 18.6 |
| 3.5 | 19.8 |
| 3.9 | 21 |
| 3.7 | 22.2 |
| 4.1 | 23.4 |
| 3.6 | 24.6 |
| 4 | 25.8 |
| 3.6 | 27 |
| 3.5 | 28.2 |
| 3.4 | 29.4 |
| 3.5 | 30.6 |
| 3.2 | 31.8 |
| 3.7 | 33 |
| 3.9 | 34.2 |
| 4.3 | 35.4 |
| 4.2 | 36.6 |
| 3.7 | 37.8 |
| 3.3 | 39 |
| 2.9 | 40.2 |
| 3.8 | 41.4 |
| 3.6 | 42.6 |
| 3.8 | 43.8 |
| 3.6 | 45 |
| 3.4 | 46.2 |

≥65 years

Incidence per 100,000

|  | Rate |
| --- | --- |
| Racialized and newcomer populations Q1 | 32.2 |
| Racialized and newcomer populations Q2 | 44.8 |
| Racialized and newcomer populations Q3 | 58.6 |
| Racialized and newcomer populations Q4 | 64.2 |
| Racialized and newcomer populations Q5 | 61 |
| Material resources Q1 | 48.8 |
| Material resources Q2 | 48.2 |
| Material resources Q3 | 46.2 |
| Material resources Q4 | 53 |
| Material resources Q5 | 60.4 |
| Households and dwellings Q1 | 37.3 |
| Households and dwellings Q2 | 40.4 |
| Households and dwellings Q3 | 40.2 |
| Households and dwellings Q4 | 49.7 |
| Households and dwellings Q5 | 76.7 |
| Age and labour Q1 | 52.1 |
| Age and labour Q2 | 56.4 |
| Age and labour Q3 | 49.9 |
| Age and labour Q4 | 45.5 |
| Age and labour Q5 | 52.2 |
| Multi-generational families Q1 | 52.3 |
| Multi-generational families Q2 | 50.3 |
| Multi-generational families Q3 | 48.9 |
| Multi-generational families Q4 | 52.5 |
| Multi-generational families Q5 | 50 |
| Unsuitable housing Q1 | 40.8 |
| Unsuitable housing Q2 | 48.2 |
| Unsuitable housing Q3 | 53.9 |
| Unsuitable housing Q4 | 65.4 |
| Average dwelling sizes Q1 | 60.4 |
| Average dwelling sizes Q2 | 44.3 |
| Average dwelling sizes Q3 | 49.1 |
| Average dwelling sizes Q4 | 53 |
| Average dwelling sizes Q5 | 45.1 |
| Household with persons under 5 years old Q1 | 54.4 |
| Household with persons under 5 years old Q2 | 51.2 |
| Household with persons under 5 years old Q3 | 47.6 |
| Household with persons under 5 years old Q4 | 47.2 |
| Household with persons under 5 years old Q5 | 52.6 |

|  |  |
| --- | --- |
| 32.2 | 0.7 |
| 44.8 | 1.8 |
| 58.6 | 3 |
| 64.2 | 4.2 |
| 61 | 5.4 |
| 48.8 | 6.6 |
| 48.2 | 7.8 |
| 46.2 | 9 |
| 53 | 10.2 |
| 60.4 | 11.4 |
| 37.3 | 12.6 |
| 40.4 | 13.8 |
| 40.2 | 15 |
| 49.7 | 16.2 |
| 76.7 | 17.4 |
| 52.1 | 18.6 |
| 56.4 | 19.8 |
| 49.9 | 21 |
| 45.5 | 22.2 |
| 52.2 | 23.4 |
| 52.3 | 24.6 |
| 50.3 | 25.8 |
| 48.9 | 27 |
| 52.5 | 28.2 |
| 50 | 29.4 |
| 40.8 | 30.6 |
| 48.2 | 31.8 |
| 53.9 | 33 |
| 65.4 | 34.2 |
| 60.4 | 35.4 |
| 44.3 | 36.6 |
| 49.1 | 37.8 |
| 53 | 39 |
| 45.1 | 40.2 |
| 54.4 | 41.4 |
| 51.2 | 42.6 |
| 47.6 | 43.8 |
| 47.2 | 45 |
| 52.6 | 46.2 |

**Figure S3**: Age-standardized annualized RSV-related hospitalization rates per 100,000 in Ontario by sociodemographic and housing characteristics, stratified by age^1^.

^1^ For each age panel, x-axis is shown on different scales.
